# Epidemiological and evolutionary considerations of SARS-CoV-2 vaccine dosing regimes

**DOI:** 10.1101/2021.02.01.21250944

**Authors:** Chadi M. Saad-Roy, Sinead E. Morris, C. Jessica E. Metcalf, Michael J. Mina, Rachel E. Baker, Jeremy Farrar, Edward C. Holmes, Oliver G. Pybus, Andrea L. Graham, Simon A. Levin, Bryan T. Grenfell, Caroline E. Wagner

## Abstract

As the threat of Covid-19 continues and in the face of vaccine dose shortages and logistical challenges, various deployment strategies are being proposed to increase population immunity levels. How timing of delivery of the second dose affects infection burden but also prospects for the evolution of viral immune escape are critical questions. Both hinge on the strength and duration (i.e. robustness) of the immune response elicited by a single dose, compared to natural and two-dose immunity. Building on an existing immuno-epidemiological model, we find that in the short-term, focusing on one dose generally decreases infections, but longer-term outcomes depend on this relative immune robustness. We then explore three scenarios of selection, evaluating how different second dose delays might drive immune escape via a build-up of partially immune individuals. Under certain scenarios, we find that a one-dose policy may increase the potential for antigenic evolution. We highlight the critical need to test viral loads and quantify immune responses after one vaccine dose, and to ramp up vaccination efforts throughout the world.

---

As the severe acute respiratory syndrome coronavirus 2 (SARS-CoV-2) betacoronavirus (*β*-CoV) pandemic continues, the deployment of safe and effective vaccines presents a key intervention for mitigating disease severity and spread and eventually relaxing non-pharmaceutical interventions (NPIs). At the time of writing, eleven vaccines have been approved. We focus on vaccines from Pfizer/BioNTech, Moderna, and Oxford/AstraZeneca. The first two elicit adaptive immunity against SARS-CoV-2 in response to the introduction of messenger ribonucleic acid (mRNA) molecules that encode the spike protein of SARS-CoV-2 (*1*), and appear to offer greater than 95% (Pfizer/BioNTech (*2*), approved in 55 countries) and 94% (Moderna (*1*), approved in 37 countries) protection against symptomatic coronavirus disease 2019 (COVID-19). Both of these mRNA vaccines were tested in clinical trials according to a two-dose regime with dose spacing of 21 and 28 days for the Pfizer/BioNTech and Moderna platforms, respectively. The Oxford/AstraZeneca vaccine uses a non-replicating adenovirus vector, and has also been tested in clinical trials according to a two-dose regime with a target 28-day inter-dose period (although for logistical reasons some trial participants received their second dose after a delay of at least 12 weeks). Clinical trials indicated 62% *-* 90% efficacy for this vaccine according to the specific dose administered (*3*). While we base our parameter choices and modeling assumptions on these three vaccines, our results are generic across platforms.

As these vaccines have been distributed internationally, several countries including the UK (*4*) and Canada (*5*) have chosen to delay the second dose in an effort to increase the number of individuals receiving at least one or in response to logistical constraints (*6*). Although a number of participants dropped out after a single dose of the vaccine in the Pfizer/BioNTech and Moderna trials, these studies were not designed to assess vaccine efficacy under these circumstances, and Pfizer has stated that there is no evidence that vaccine protection from a single dose extends beyond 21 days (*4*). The Oxford/AstraZeneca clinical trials did include different dose spacings, and limited evidence suggests that longer intervals (two to three months) did not affect and may even have improved vaccine efficacy (*3, 4*). Ultimately, the consequences of deviating from manufacturer-prescribed dosing regimes at the population scale remain unknown, but will hinge on immune responses.

While there has been significant progress in quantifying host immune responses following infection (*7*), substantial uncertainty regarding the strength and duration of both natural and vaccinal SARS-CoV-2 immunity remains. Previous work suggests that these factors will play a central role in shaping the future dynamics of Covid-19 cases (*8*). Future cases also create an environment for the selection of novel variants (e.g. (*9–11*)). Of particular concern is the possibility of antigenic drift (via immune escape from natural or vaccinal immunity), especially if immunity elicited after a single vaccine dose is weaker than that of the complete two-dose regime. Consequently, the longer term epidemiological and evolutionary implications of these different SARS-CoV-2 vaccine dosing regimes are not yet clear; the immediate need for effective mass vaccination makes understanding them critical to inform policy.

Here, we explore these epidemiological and evolutionary considerations with an extension of a recent immuno-epidemiological model for SARS-CoV-2 dynamics (*8*), depicted schematically in Figure 1. Without vaccination, our model reduces to the Susceptible-Infected-Recovered-(Susceptible) (SIR(S)) model (*8, 12*), where individual immunity after recovery from primary infection may eventually wane, leading to potentially reduced susceptibility to secondary infections, denoted by the fraction *ϵ* relative to a baseline level of unity. This parameter *ϵ* thus titrates between the SIR (lifetime immunity, *ϵ* = 0) and SIRS (hosts regain complete susceptibility, *ϵ* = 1) paradigms. In this model extension (Fig. 1 and Supplementary Materials) we incorporate two vaccinated classes; *V*_1_ accounts for individuals who have received one dose of a SARS-CoV-2 vaccine and *V*_2_ tracks individuals who have received two doses. In the short term, we assume that both dosing options decrease susceptibility by fractions 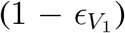 (one dose) and 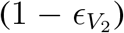 (two doses), inferred from the clinical trial data (though the nature of the infecting variant may influence this); we also assume that *I*_*V*_ tracks infection following vaccination. We allow for vaccinal immunity to wane at separate rates (*ρ*_1_ (one dose) and *ρ*_2_ (two doses)), moving individuals to the partially susceptible immune classes 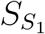 and 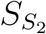 characterized by (possibly different) levels of immune protection *ϵ*_1_ and *ϵ*_2_. Infection following waned one-dose or two-dose vaccinal immunity is tracked by the immune classes 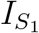 and 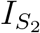, respectively. We consider a continuous spectrum for the inter-dose period 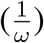 with an infinite value corresponding to a “one-dose strategy”, and model the rate of administration of the first dose *ν* as an increasing function of the inter-dose period (Fig. 1 and Supplementary Materials) to reflect the increase in available doses due to a delayed second dose. Thus, dosing regimes with longer inter-dose periods allow for higher coverage with the first dose.

**Figure 1:**
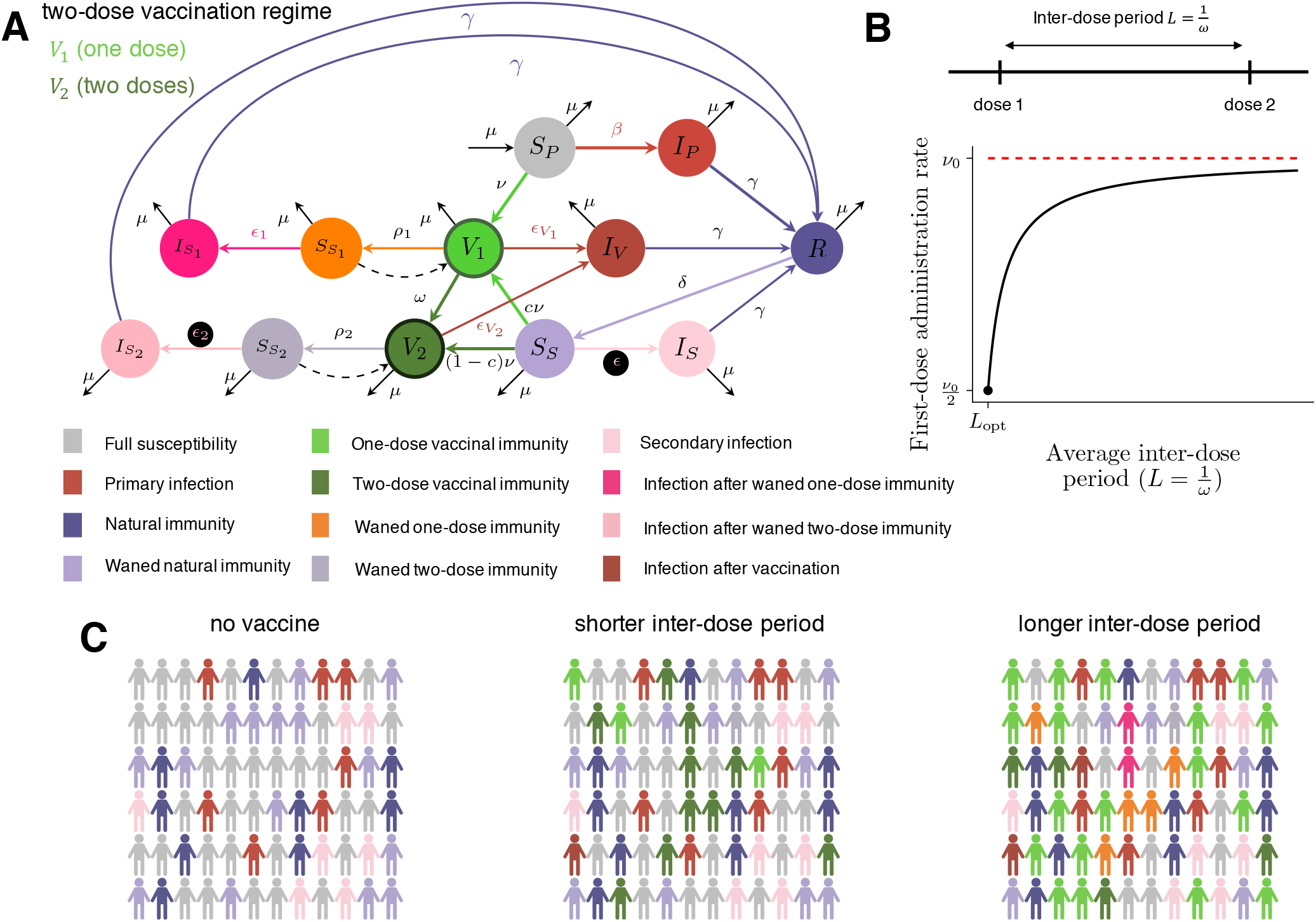
Description of the extended immuno-epidemiological model with one- and two-dose vaccination regimes (based on (*8*)). (A) Model flow chart depicting transitions between immune classes (see main text and Supplementary Materials for a full description of the immune classes and parameters). (B) Diagram of the inter-dose period 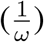 considered between the first and second vaccine doses and its relationship to the rate of administration of the first vaccine dose *ν*. The maximum achievable rate is *ν*_0_ for a fully one-dose strategy, and *ν* is assumed to decrease exponentially to its lowest value *ν*_0_/2 when a fully two-dose strategy with inter-dose period corresponding to the clinical recommendation *(L*_*opt*_) is employed. (C) Representative schematic of societal composition of various immune classes for the SIR(S) model with no vaccination (left), the extended model with a short inter-dose period (middle), and the extended model with a long inter-dose period (right).

We begin our analysis by studying the epidemiological impacts of the different dosing regimes on the medium-term temporal dynamics of Covid-19 cases. We then examine the potential evolutionary consequences of dosing regime through the quantification of a time-dependent relative net viral adaptation rate (*13*). This term is related to the strength of the conferred natural and vaccinal immunity (via either inducing selection through immune pressure or suppressing viral replication) as well as the sizes of classes of individuals experiencing infections after immune waning.

## Epidemiological impacts

As a base case, we consider a high latitude European or North American city with initial conditions that qualitatively correspond to early 2021 (see Supplementary Materials and Figures S5 and S6 for other scenarios, e.g. a high initial attack rate or almost full susceptibility), in addition to a seasonal transmission rate (*16*) with NPIs (see Supplementary Materials). Furthermore, the UK and Canadian policy is for a delayed second dose; they are not aiming for an “exclusively” one-dose policy. However, we explore the one-dose strategy as an extreme case for the ‘two-dose’ vaccines; it also encompasses a pessimistic situation of waning public opinion on vaccination and individuals’ own decisions to forgo the second dose. Finally, this one-dose policy could capture vaccines which only require a single dose, e.g. the Johnson & Johnson vaccine.

In Figure 2, we present potential scenarios for medium-term SARS-CoV-2 infection and immunity dynamics contingent upon vaccine dosing regimes. We start by assuming that vaccination occurs at a constant rate, and assume a relatively optimistic maximum rate of administration of the first dose of *ν*_0_ = 2% of the population per week (see Supplementary Materials for other scenarios). Figures 2A and 2B correspond, respectively, to scenarios with weaker (and shorter) and stronger (and longer) natural and vaccinal adaptive immune responses. Thus, the former represents a scenario with higher secondary susceptible density than the latter. In each panel, the top and bottom sections consider poor and robust one-dose vaccinal immunity, respectively. The leftmost column represents a one-dose vaccine policy (captured in the model by infinite dose spacing), with dose spacing decreasing to 4 weeks in the rightmost column (i.e. a strict two-dose policy with doses separated by the clinical trial window corresponding to Moderna’s recommendations for their vaccine, hereafter referred to as the “recommended two-dose strategy”).

**Figure 2:**
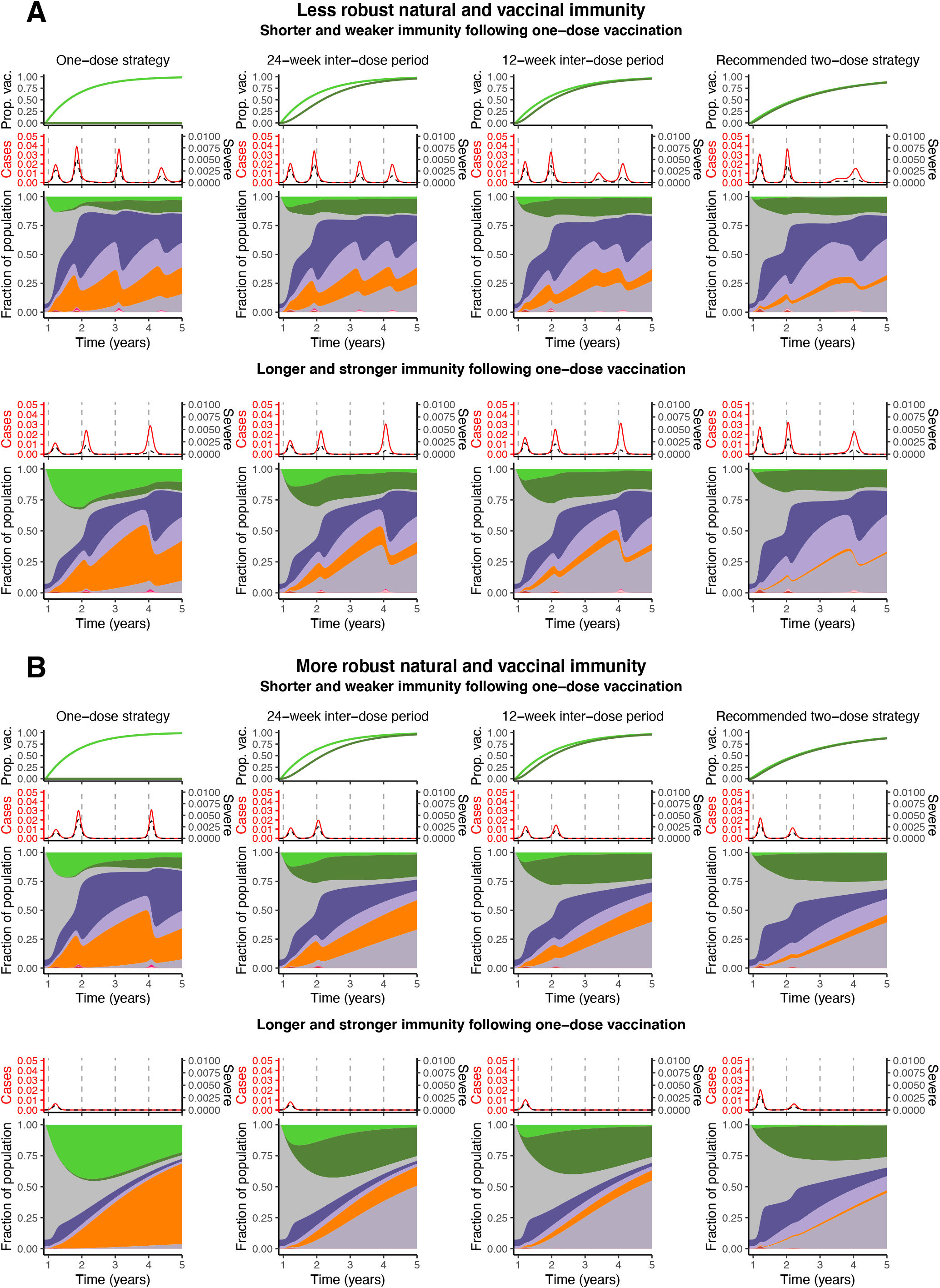
Illustrative time series of the fraction of the population vaccinated with one or two doses (top) (see note (*14*)), the fraction of total and severe infections (see (*15*)) (middle), and area plots of the fraction of the population comprising each immune 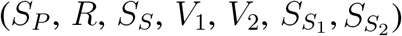 infection 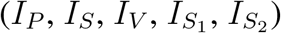 class (bottom) from the introduction of vaccination until 5 years after the pandemic onset. The immune and infection class colors are the same as those defined in Figure 1A. In all plots, the maximum rate of administration of the first vaccine dose is taken to be *ν*_0_ = 2% and the vaccine is introduced at *t*_vax_ = 48 weeks. We take 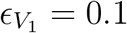 and 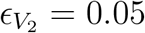 in keeping with data from clinical trials (*2*). The fraction of severe cases for primary infections, secondary infections, infection after vaccination, and infection after waned two-dose immunity are taken to be *x*_sev,p_ = 0.14, *x*_sev,s_ = 0.07, *x*_sev,V_ = 0.14, and *x*_sev,2_ = 0. The transmission rates and periods of NPI adoption are defined in the Supplementary Materials. The leftmost column corresponds to a one-dose vaccine strategy (*ω* = 0), followed by inter dose spacings of 24 weeks, 12 weeks, and 4 weeks (rightmost column). (A) corresponds to an overall more pessimistic natural and vaccinal immunity scenario, with *ϵ* = *ϵ*_2_ = 0.7 and 1/*δ* = 1/*ρ*_2_ = 1 year. For a less effective one-dose vaccine (top section), we take *ϵ*_1_ = 0.9, 1/*ρ*_1_ = 0.25 years, and the fraction of severe cases associated with infection after waned one-dose immunity is *x*_sev,1_ = 0.14. For an effective one-dose vaccine (bottom section), we take *ϵ*_1_ = 0.7, 1/*ρ*_1_ = 1 year, and the fraction of severe cases associated with infection after waned one-dose immunity is *x*_sev,1_ = 0. (B) corresponds to an overall more optimistic natural and vaccinal immunity scenario, with *ϵ* = *ϵ*_2_ = 0.5 and 1/*δ* = 1/*ρ*_2_ = 2 years. For a less effective one-dose vaccine (top section), we take *ϵ*_1_ = 0.9, 1/*ρ*_1_ = 0.5 years, and the fraction of severe cases associated with infection after waned one-dose immunity is *x*_sev,1_ = 0.14. For an effective one-dose vaccine (bottom section), we take *ϵ*_1_ = 0.5, 1/*ρ*_1_ = 2 years, and the fraction of severe cases associated with infection after waned one-dose immunity is *x*_sev,1_ = 0.

As expected, we find that broader deployment of widely-spaced doses is beneficial. Specifically, a one-dose strategy (or a longer inter-dose period) may lead to a substantially reduced ‘first’ epidemic peak of cases after the initiation of vaccination (compare the leftmost top panels of Figs. 2A and 2B with the no vaccination scenarios in Figs. S1A and S1B). This result applies even if immunity conferred by one vaccine dose is shorter and weaker than that following two-doses (top panels of Figures 2A and 2B). However under these conditions of imperfect immunity, an exclusively one-dose strategy then leads to an earlier subsequent peak due to the accumulation of partially susceptible individuals. When the rate of administration of the first dose is very high (Fig S4, *ν*_0_ = 5% per week), this subsequent infection peak may be larger than that expected in the scenario with no vaccination. In general, the accumulation of partially susceptible individuals with waned one-dose vaccinal immunity can be mitigated by implementing a two-dose strategy and decreasing the time between doses. Thus, in situations of a less effective first dose where the second dose is delayed, it is important to ensure individuals eventually do obtain their second dose.

In line with intuition, longer and stronger immunity elicited after a single dose heightens the benefits of a one-dose strategy or of delaying the second dose (compare the top and bottom left-most panels of Figs. 2A and 2B). Additionally, the protective effects of adopting these strategies instead of the two-dose regime are maintained in the medium-term, with decreased burden in all future peaks. This is further summarized in Figure 3A, where the cumulative number of total and severe cases (right and left panels, respectively), from the time of vaccine initiation through the end of the five year period considered normalized by the burdens with no vaccination, are plotted as a function of the inter dose period and the one- to two-dose immune response ratio *x*_*e*_ (see figure caption for details). When the immune response conferred by a single dose is nearly or as robust as that following two doses, total case numbers (Figure 3A, right panel) can be substantially reduced by delaying the second dose. However, for smaller values of *x*_*e*_, larger inter-dose periods are associated with more cases. The reduction in the cumulative burden of severe cases is even more sizeable (Figure 3A, left panel) due to the assumed reduction in the fraction of severe cases for partially immune individuals. When vaccination rates are substantially lower (Fig S2, *ν*_0_ = 0.1% per week and Fig S3, *ν*_0_ = 1% per week), the benefits of a single dose strategy diminish even for an effective first dose, as an insufficient proportion of the population are immunized. The effect of the vaccine on case numbers is sensitive to when it is introduced in the dynamical cycle (Figs. S7, S8), highlighting the critical interplay between the force of infection and the level of population immunity (see Supplementary Materials for further details).

**Figure 3:**
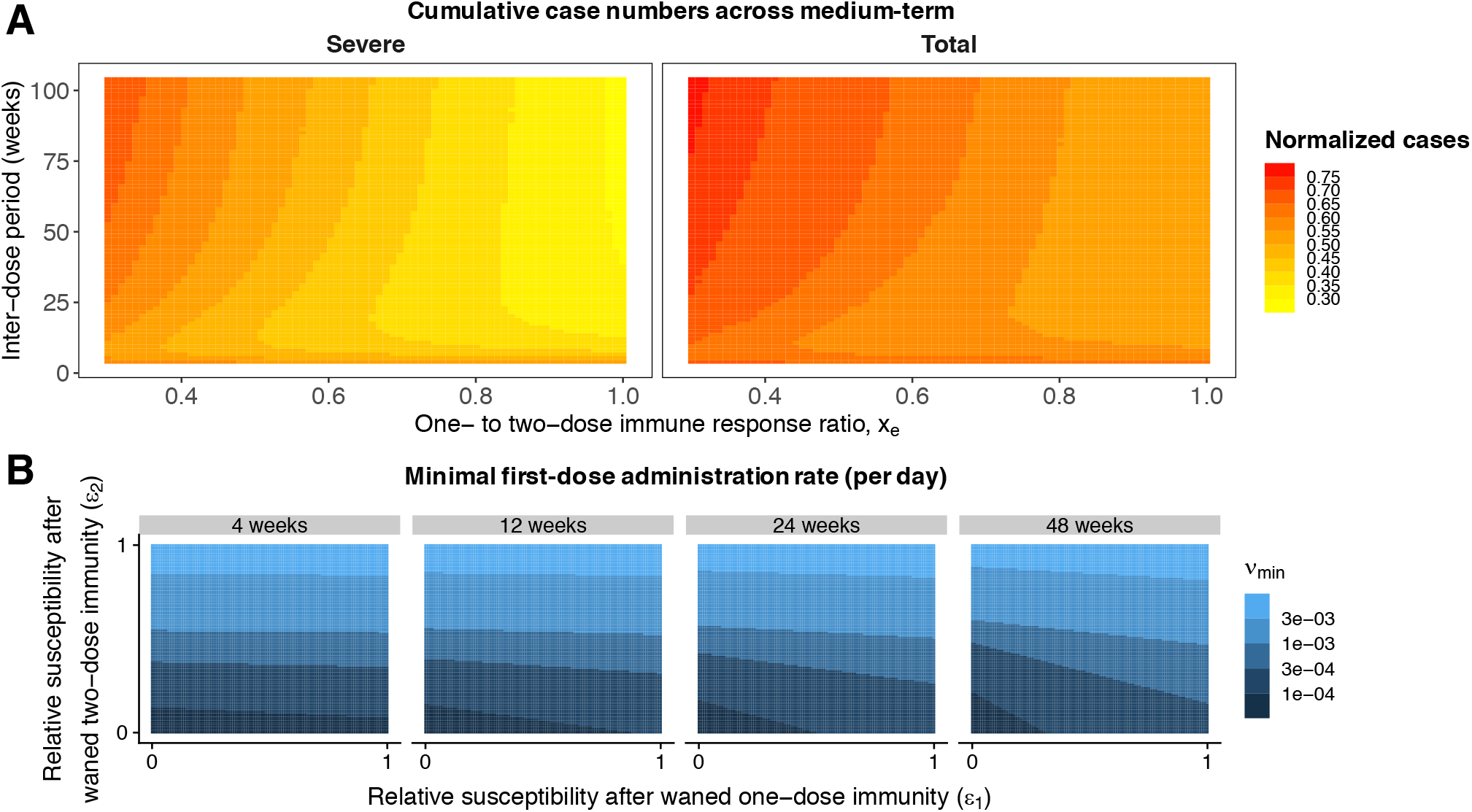
Heat maps depicting various epidemiological outcomes contingent on dosing regimes. (A) Cumulative severe (left) and total (right) case numbers relative to the scenario with no vaccine from the time of vaccine introduction through the end of the five-year time period following the onset of the pandemic as a function of the one- to two-dose immune response ratio *x*_*e*_ and the inter-dose period. Parameters correspond to the “weak” immunity scenario of Figure 2A, but *x*_*e*_ sets the value of *ϵ*_1_, *ρ*_1_, and *x*_sev,1_. Specifically, we take *ϵ*_1_ = *ϵ*_2_ + (1 − *x*_*e*_)(1 − *ϵ*_2_) such that the susceptibility to infection after a waned single dose interpolates linearly between the value after waned two doses (*ϵ*_2_) when the one and two dose immune responses are equally strong (*x*_*e*_ = 1) and unity (full susceptibility) when a single dose offers no immune protection (*x*_*e*_ = 0). Similarly, we take *x*_sev,1_ = *x*_sev,2_ +(1 − *x*_*e*_)(*x*_sev,V_ − *x*_sev,2_) such that the fraction of severe cases for infections following a waned single dose interpolates linearly between the value after waned two doses (*x*_sev,2_) when *x*_*e*_ = 1 and the value after a (failed) vaccination *x*_sev,V_ when *x*_*e*_ = 0. Finally, *ρ*_1_ is given by *ρ*_1_ = *ρ*_2_/x_*e*_. (B) Values of *ν*_min_, the minimal rate of first dose administration per day such that for any *ν* > *ν*_min_ the basic reproduction ℛ_0_[*ν*] < 1 and the disease cannot invade (see Supplementary Materials), as a function of the strength of immunity following one (*ϵ*_1_) and two (*ϵ*_2_) waned vaccines doses, for different inter-dose periods. We take the duration of one dose and two dose vaccinal immunity to be 1/*ρ*_1_ = 0.5 years and 1/*ρ*_2_ =1 year, respectively, and set 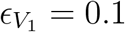 and 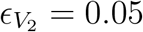.

Vaccinal immunity will be central to efforts to attain community immunity and prevent local spread due to case importation. We therefore analytically calculated the first vaccine dose administration rate for a given inter-dose spacing required for community immunity in our model (see Supplementary Materials). In the long term, however, individuals whose one- or two-dose immunity has waned will likely be able to be vaccinated again before infection, and so we incorporated re-vaccination of these individuals into the extended model and computed an analogous minimal vaccination rate which we plot in Figure 3B. We find that as the inter-dose period grows, this minimal rate depends increasingly on the degree of reduction in susceptibility after the waning of one-dose vaccinal immunity *ϵ*_1_ (Figure 3B and see Figure S13 for other parameter choices). Vaccine refusal (*17*) may also impact the attainment of community immunity through vaccinal immunity in the longer-term (see Supplementary Materials).

While we have assumed that the inter-dose period is exponentially distributed, we have relaxed this assumption and examined an Erlang-distributed inter-dose period (see Supplementary Materials). The model predictions are qualitatively and quantitatively similar (compare Figure 2 with Fig. S9), justifying our choice of the simpler model.

## Evolutionary impacts

The recent emergence of numerous SARS-CoV-2 variants in still relatively susceptible populations underline the virus’s evolutionary potential (*18–20*). We focus here on the longer term potential for immune escape from natural or vaccinal immunity (*13*). For immune escape variants to spread within a population, they must first arise via mutation, and then there must be substantial selection pressure in their favour. We expect the greatest opportunity for variants to arise in (and spread from) hosts with the highest viral loads, likely those with the least immunity. On the other hand, we expect the greatest selection where immunity is the greatest. Previous research on the phylodynamic interaction between viral epidemiology and evolution (based on seasonal influenza) predicts that partially immune individuals (permitting intermediate levels of selection and transmission) could maximize levels of escape ((*13*), Figure 4A). This is consistent with case reports of sustained antigenic evolution in immunocompromised patients with prolonged Covid-19 infections (*21*). Under this model, we would project that different categories of secondarily infected people (after waning of natural immunity or immunity conferred from one or two doses of vaccine) would be key potential contributors to viral immune escape.

**Figure 4:**
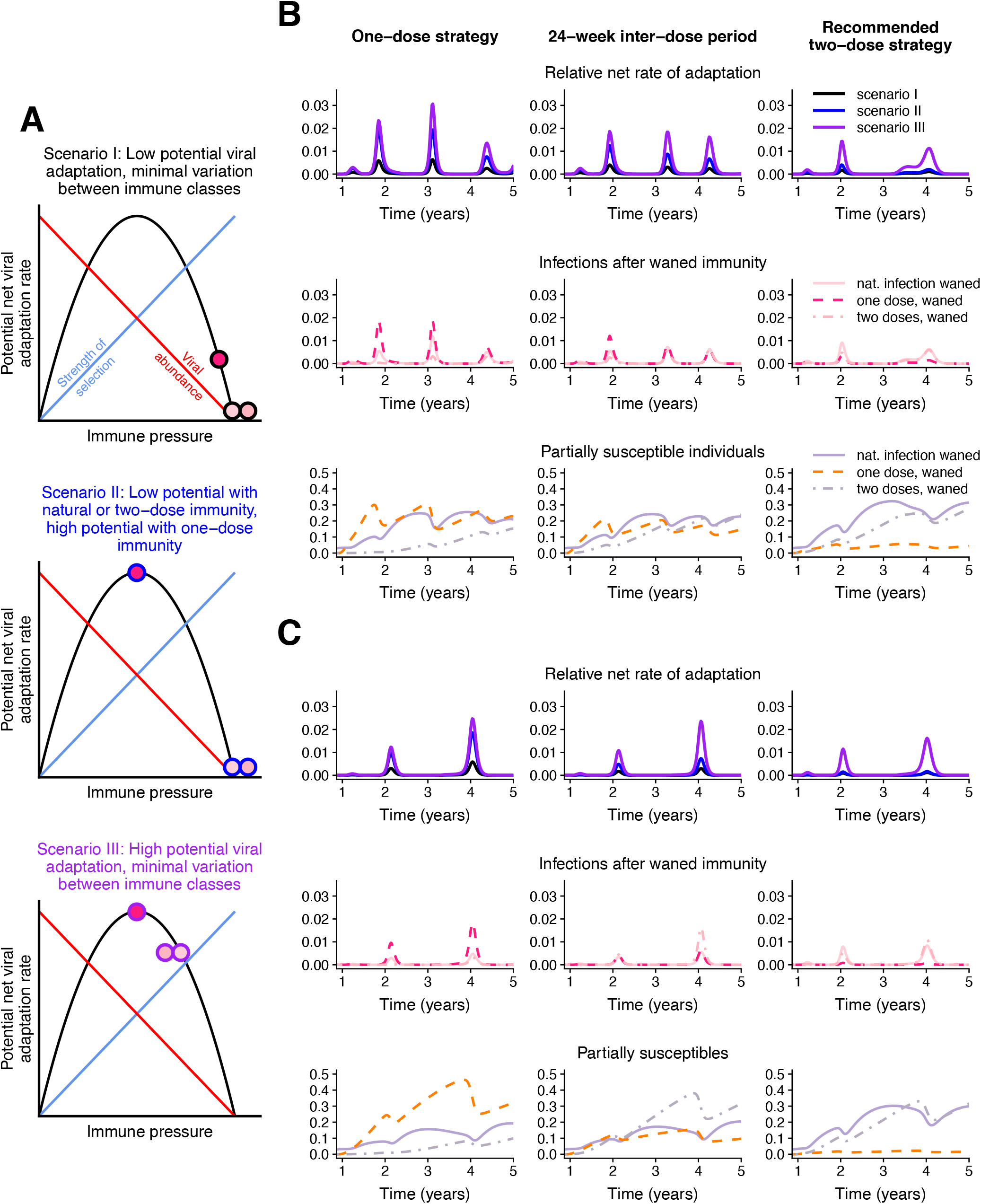
Potential viral evolution scenarios under different vaccine regimes. (A) Schematic representations of the potential net viral adaptation rate associated with the *I*_*S*_, 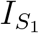, and 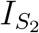 infection classes under three different scenarios. These are illustrated by the filled dots, with the central colour denoting the infection class and corresponding to the legend in Figure 1A. The dot outlines correspond to the three scenarios considered (Scenario I: black lines and top panel, Scenario II: blue lines and middle panel, and Scenario III: purple lines and bottom panel). The phylodynamic model for potential viral adaptation as a function of immune pressure is adapted from (*13*). (B) and (C): relative net rates of adaptation (top rows; colours correspond to the scenarios in (A)), and composition of associated infection (*I*_*S*_: solid lines, 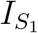: dashed lines, 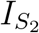: dashed-dotted lines; middle rows) and susceptible (*S*_*S*_: solid lines, 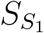: dashed lines, 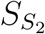: dashed-dotted lines; bottom rows) classes. The colours in the middle and bottom rows correspond to the legend in Figure 1A. The leftmost column corresponds to a one dose strategy, an inter-dose period of 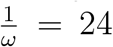 weeks is assumed in the middle column, and the rightmost column assumes a two dose strategy with doses separated by the recommended window of 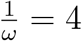 weeks. Both (B) and (C) correspond to a “weak” natural and vaccinal immunity scenario, with the same parameters as those in Figure 2A. A weaker immune response after one vaccine dose is assumed in (B) (with parameters corresponding to those in the top section of Figure 2A), and a stronger immune response after one vaccine dose is assumed in (C) (with parameters corresponding to those in the bottom section of Figure 2A). The weights used to calculate the relative net rates of adaptation are *w*_*IS,I*_ = 0.05, *w*_*IS1,I*_ = 0.3, and *w*_*IS2,I*_ = 0.05 in Scenario I, *w*_*IS,II*_ = 0.05, *w*_*IS1,II*_ = 1, and *w*_*IS2,II*_ = 0.05 in Scenario II, and *w*_*IS,III*_ = 0.8, *w*_*IS1,III*_ = 1, and *w*_*IS2,III*_ = 0.8 in Scenario III.

In Figure 4, we explore three potential evolutionary scenarios, each with their own assumptions regarding viral abundance and within-host selection for the different immune classes. In all scenarios, we assume for simplicity that immunity elicited after two doses of the vaccine is equivalent to that elicited after natural infection. We also assume that transmission rises with viral abundance in hosts (*13*). In Scenario I (black borders on circles, top panel of Figure 4A), we assume that infections of all classes of partially susceptible individuals lead to strong selective pressures and low viral abundance (a marker of low transmission), and thus low rates of adaptation, with only slightly reduced immune pressure for infections after a waned single vaccine dose relative to natural infection or two doses. Scenario II (blue borders on circles, middle panel of Figure 4A), considers a situation where natural and two-dose vaccinal immunity again lead to low viral abundance, but one-dose vaccinal immunity is associated with intermediate immune pressure that results in substantially higher rates of viral adaptation. Finally, in Scenario III (purple borders on circles, bottom panel of Figure 4A), adaptive immune responses following waned natural, one dose, and two dose vaccinal immunity all lead to similar intermediate levels of immune pressure and high rates of viral adaptation. In all cases, we assume for tractability that viral immune escape is not correlated with clinical severity (*22*).

The relative potential viral adaptation rates (see (*13*) for more details) corresponding to each scenario are presented in the top rows of Figures 4B and 4C. This relative rate is estimated as the sum of the sizes of the infection classes following waned immunity (i.e. *I*_*S*_ after 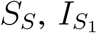 after 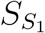, and 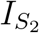 after 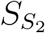) weighted by the infection class-specific net viral adaptation rate assigned in each scenario. Therefore, this quantity reflects a weight-averaged potential rate for viral adaptation per-individual per-infection. The corresponding immune and susceptibility classes are plotted in the middle and bottom rows, respectively, according to the colour scheme defined in Figure 1A. The weaker immunity scenario of Figure 2A is considered, with Figures 4B and 4C corresponding, respectively, to the situations of a weaker and more robust single vaccine dose relative to two doses. The leftmost column corresponds to a one dose strategy, an inter-dose period of 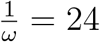 weeks is assumed in the middle column, and the rightmost column assumes a two dose strategy with doses separated by the clinical trial window of 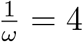 weeks.

Different assumptions regarding the strength and duration of adaptive immune responses to vaccines and natural infections result in different predictions for the proportions of individuals in the partially susceptible immune classes over time. When one dose vaccinal immunity is poor, a one-dose strategy results in the rapid accumulation of partially susceptible 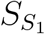 individuals (Figure 4B, bottom row) and a greater infection burden. When the assumed individual rates of evolutionary adaptation arising from these infection classes are high (Scenarios II and III), we find that a one-dose strategy could lead to substantially higher relative rates of adaptation. This effect can be mitigated by implementing a two-dose strategy even with a longer inter-dose period than the recommended duration, echoeing our epidemiological findings.

When one dose vaccinal immunity is strong, reduced infection burdens result in lower relative rates of adaptation when a one dose strategy is used, although the large fraction of 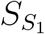 individuals may still lead to evolutionary pressure, particularly when the potential viral adaptation rate associated 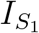 infections is large. A two-dose strategy mitigates this effect, but the corresponding reduction in vaccinated individuals increases the infection burden from other classes. Thus, to avoid these potentially pessimistic evolutionary outcomes, our results highlight the importance of rapid vaccine deployment. More broadly, our results further underline the importance of equitable, global vaccine deployment (*23, 24*): immune escape anywhere will quickly spread.

## Impact of increasing vaccination through time

In Supplementary Materials (Figures S10, S11, S12), we explore the implications of ramping up vaccine deployment through two approaches. First, we examine a simple increase in the rate of administration of the first dose and unchanged dosing regimes (Fig. S10). Qualitatively, these results are largely analogous to our previous results, and reflect the benefits of increasing population immunity through an increase in vaccination deployment.

However, as vaccines become more widely available, policies on dosing regimes may change. The second approach we consider is a timely shift to a two-dose policy with recommended interdose spacing as vaccine deployment capacity increases (Figs. S11, S12). Initially delaying (or omitting) the second dose decreases the first epidemic peak after the initiation of vaccination. Such a reduction in first peak size would also reduce secondary infections, and thus potentially immune escape in most cases (i.e. an evolutionary advantage). Subsequently, the switch to a manufacturer-timed vaccine dosage regime mitigates the potential medium-term disadvantages of delaying (or omitting) the second dose that may arise if immunity conferred from a single dose is relatively poor, including the accumulation of partially susceptible 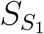 individuals whose one-dose vaccinal immunity has waned. These contrasts highlight the importance of data-driven policies that undergo constant re-evaluation as vaccination progresses.

## Caveats

Our immuno-epidemiological model makes several assumptions. While heterogeneities (super-spreading, age, space, etc) (*25–27*) are important for the quantitative prediction of SARS-CoV-2 dynamics, we previously found that these do not qualitatively affect our results (*8*). Nevertheless, we again briefly explore heterogeneities in transmission and vaccine coverage in the Supplementary Materials. We have also assumed that the robustness of immune responses following the second dose is independent of the inter-dose period, yet it is possible that delaying the second dose may actually enhance adaptive immune responses. Detailed clinical evaluation of adaptive immune responses after one and two vaccine doses with different inter-dose spacing is an important direction for future work.

Additionally, we have assumed highly simplified scenarios for NPIs. The chosen scenario was selected to qualitatively capture current estimates of SARS-CoV-2 prevalence and seropositivity in large cities. However, these values vary substantially between locations, a notable example being recent estimates of a large infection rate in Manaus, Brazil during the first wave (*28*), or countries having almost no infections due to the successful implementation of NPIs (*29–31*). We have examined these scenarios in the Supplementary Materials (Figures S5 and S6). The qualitative projections of our model are sensitive to the composition of infection and immune classes at the onset of vaccination (including, therefore, the assumption of dramatically higher seropositivity levels, i.e. the sum of the *S*_*S*_ and *R* classes). We further explore this in the Supplementary Materials through the initiation of vaccination at different times in the dynamic cycle (Figs. S7 and S8). Thorough explorations of various NPIs, seasonal transmission rate patterns, vaccine deployment rates, dosing regimes, and clinical burdens will be able to be investigated for broad ranges of epidemiological and immunological parameters with an online interactive application upon publication.

Finally, we have explored the simplest evolutionary model, which can only give a general indication of the potential for evolution under different scenarios. Including more complex evolutionary models (*32, 33*) into our framework is thus another important area for future work. A full list of caveats is presented in Supplementary Materials.

## Conclusion

The deployment of SARS-CoV-2 vaccines in the coming months will strongly shape post-pandemic epidemiological trajectories and characteristics of accumulated population immunity. Dosing regimes should seek to navigate existing immunological and epidemiological trade-offs between individuals and populations. Using simple models, we have shown that different regimes may have crucial epidemiological and evolutionary impacts, resulting in a wide range of potential outcomes in the medium term. Our work also lays the foundation for a number of future considerations related to vaccine deployment during ongoing epidemics, especially preparing against future pandemics.

In line with intuition, spreading single doses in emergency settings (i.e. rising infections) is beneficial in the short term and reduces prevalence. Furthermore, we find that if immunity following a single dose is robust, then delaying the second dose is also optimal from an epidemiological perspective in the longer term. On the other hand, if one-dose vaccinal immunity is weak, the outcome could be more pessimistic; specifically, a vaccine strategy with a very long inter-dose period could lead to marginal short-term benefits (a decrease in the short-term burden) at the cost of a higher infection burden in the long term and substantially more potential for viral evolution. These negative longer term effects may be alleviated by the eventual administration of a second dose, even if it is moderately delayed. With additional knowledge of the relative strength and duration of one-dose vaccinal immunity and corresponding, clinically-informed policies related to dosing regimes, pessimistic scenarios may be avoided.

In places where vaccine deployment is delayed and vaccination rates are low, our results stress the subsequent negative epidemiological and evolutionary impacts that may emerge. Particularly since these consequences (e.g., the evolution of new variants) could emerge as global problems, there is an urgent need for global equity in vaccine distribution and deployment (*23, 24*).

Current uncertainties surrounding the strength and duration of adaptive immunity in response to natural infection or vaccination lead to very broad ranges for the possible outcomes of various dosing regimes. Nevertheless, ongoing elevated Covid-19 case numbers stresses the rapid need for effective, mass vaccine deployment. Overall, our work emphasizes that the impact of vaccine dosing regimes are strongly dependent on the relative robustness of immunity conferred by a single dose. It is therefore imperative to determine the strength and duration of clinical protection and transmission-blocking immunity through careful clinical evaluations (including, for instance, randomized control trials of dose intervals and regular testing of viral loads in vaccinated individuals, their contacts, and those who have recovered from natural infections) in order to enforce sound public policies. Our results underscore the importance of exploring the phylodynamic interaction of pathogen dynamics and evolution, from within host to global scales, for SARS-CoV-2, influenza, and other important pathogens (*32–37*).

## Data Availability

The manuscript does not include new data

## Supplementary Materials

The Supplementary Materials contain technical details, expanded analyses, supplementary figures, and references. See attached document.

## Acknowledgements

This work was funded in part by Open Philanthropy, the Natural Sciences and Engineering Research Council of Canada through a Postgraduate-Doctoral Scholarship (CMSR), the Co-operative Institute for Modelling the Earth System (CIMES) (REB), the James S. McDonnell Foundation 21st Century Science Initiative Collaborative Award in Understanding Dynamic and Multi-scale Systems (CMSR, SAL), the C3.ai Digital Transformation Institute and Microsoft Corporation (SAL), Gift from Google, LLC (SAL), the National Science Foundation (CNS-2027908, CCF1917819) (SAL), the U.S. CDC (BTG), Flu Lab (BTG).

