## Supplementary Material for "Epidemiological and evolutionary considerations of SARS-CoV-2 vaccine dosing regimes"

### Contents

**Model formulation** **3**

**Determination of seasonal reproduction numbers** **4**

**Modeling of nonpharmaceutical interventions (NPIs)** **4**

**Linking vaccination rate to inter-dose period** **5**

**Calculation of the basic reproduction number $\mathcal{R}_0$** **5**

**Vaccine refusal** **9**

**Other heterogeneities in vaccination deployment** **11**

**Distribution of inter-dose period** **11**

**Additional Scenarios** **12**

**Caveats and future directions** **15**

**Supplementary figures** **17**

**References** **30**

### Model formulation

We extend the model of (*1*) to examine different vaccination strategies. The additional compartments are as follows: $V_i$ denotes individuals vaccinated with $i$ doses and are thus immune; $S_{S_i}$ denotes individuals whose complete $i$-dose immunity has waned and are now partially susceptible again; $I_{S_i}$ denotes individuals who were in $S_{S_i}$ and have now been infected again; $I_V$ denotes individuals for whom the vaccine did not prevent infection.

The extended model contains several new parameters: $\frac{1}{\rho_i}$ is the average duration of vaccinal immunity $V_i$; $\frac{1}{\omega}$ is the average inter-dose period; $\epsilon_{V_i}$ is the decrease in susceptibility following vaccination with dose $i$; $\epsilon_i$ is the decrease in susceptibility following waning of $i$-dose immunity; $\alpha_i$ is the relative infectiousness of individuals in $I_{S_i}$; and $\alpha_V$ is the relative infectiousness of individuals in $I_V$. To allow for heterogeneity in vaccinal immune responses and potentially cumulative effects of natural and vaccinal immunity, we take $c$ to be the fraction of previously-infected partially susceptible individuals ($S_S$) for whom one dose of the vaccine gives equivalent immunity to two-doses for fully susceptible individuals ($S_P$). Finally, $x_i$ is the fraction of individuals in $S_{S_i}$ that are re-vaccinated, and (1-$p_i$) is the fraction of individuals in $S_{S_i}$ for whom re-administration of the "first dose" provides equivalent immune protection to two doses (i.e. they transition to the $V_2$ class). The full set of equations governing the transitions between these infection and immunity classes is then given by:

$$\frac{dS_P}{dt} = \mu - \beta S_P[I_P + \alpha I_S + \alpha_V I_V + \alpha_1 I_{S_1} + \alpha_2 I_{S_2}] - (s_{\text{vax}}\nu + \mu)S_P, \tag{S1a}$$

$$\frac{dI_P}{dt} = \beta S_P[I_P + \alpha I_S + \alpha_V I_V + \alpha_1 I_{S_1} + \alpha_2 I_{S_2}] - (\gamma + \mu)I_P, \tag{S1b}$$

$$\frac{dR}{dt} = \gamma(I_P + I_S + I_V + I_{S_1} + I_{S_2}) - (\delta + \mu)R, \tag{S1c}$$

$$\frac{dS_S}{dt} = \delta R - \epsilon\beta S_S[I_P + \alpha I_S + \alpha_V I_V + \alpha_1 I_{S_1} + \alpha_2 I_{S_2}] - (s_{\text{vax}}\nu + \mu)S_S, \tag{S1d}$$

$$\frac{dI_S}{dt} = \epsilon\beta S_S[I_P + \alpha I_S + \alpha_V I_V + \alpha_1 I_{S_1} + \alpha_2 I_{S_2}] - (\gamma + \mu)I_S, \tag{S1e}$$

$$\begin{aligned}\frac{dV_1}{dt} &= s_{\text{vax}}\nu S_P + c s_{\text{vax}}\nu S_S + x_1 p_1 s_{\text{vax}}\nu S_{S_1} + x_2 p_2 s_{\text{vax}}\nu S_{S_2} \\ &\quad - \epsilon_{V_1}\beta V_1[I_P + \alpha I_S + \alpha_V I_V + \alpha_1 I_{S_1} + \alpha_2 I_{S_2}] - (\omega + \rho_1 + \mu)V_1,\end{aligned} \tag{S1f}$$

$$\begin{aligned}\frac{dV_2}{dt} &= (1-c)s_{\text{vax}}\nu S_S + x_1(1-p_1)s_{\text{vax}}\nu S_{S_1} + x_2(1-p_2)s_{\text{vax}}\nu S_{S_2} + \omega V_1 \\ &\quad - \epsilon_{V_2}\beta V_2[I_P + \alpha I_S + \alpha_V I_V + \alpha_1 I_{S_1} + \alpha_2 I_{S_2}] - (\rho_2 + \mu)V_2,\end{aligned} \tag{S1g}$$

$$\frac{dI_V}{dt} = \beta(\epsilon_{V_1}V_1 + \epsilon_{V_2}V_2)[I_P + \alpha I_S + \alpha_V I_V + \alpha_1 I_{S_1} + \alpha_2 I_{S_2}] - (\gamma + \mu)I_V, \tag{S1h}$$

$$\frac{dS_{S_1}}{dt} = \rho_1 V_1 - \epsilon_1\beta S_{S_1}[I_P + \alpha I_S + \alpha_V I_V + \alpha_1 I_{S_1} + \alpha_2 I_{S_2}] - (s_{\text{vax}}x_1\nu + \mu)S_{S_1}, \tag{S1i}$$

$$\frac{dS_{S_2}}{dt} = \rho_2 V_2 - \epsilon_2 \beta S_{S_2}[I_P + \alpha I_S + \alpha_V I_V + \alpha_1 I_{S_1} + \alpha_2 I_{S_2}] - (s_{\text{vax}} x_2 \nu + \mu) S_{S_2}, \tag{S1j}$$
$$\frac{dI_{S_1}}{dt} = \epsilon_1 \beta S_{S_1}[I_P + \alpha I_S + \alpha_V I_V + \alpha_1 I_{S_1} + \alpha_2 I_{S_2}] - (\gamma + \mu) I_{S_1}, \tag{S1k}$$
$$\frac{dI_{S_2}}{dt} = \epsilon_2 \beta S_{S_2}[I_P + \alpha I_S + \alpha_V I_V + \alpha_1 I_{S_1} + \alpha_2 I_{S_2}] - (\gamma + \mu) I_{S_2}. \tag{S1l}$$

For all simulations, we take $\mu = 0.02\text{y}^{-1}$ corresponding to a yearly crude birth rate of 20 per $1000$ people. Additionally, we take the infectious period to be $1/\gamma = 5$ days, consistent with the modeling in (*1–3*) and the estimation of a serial interval of $5.1$ days for Covid-19 in (*4*), and assume that $c = 0.5$. We take the relative transmissibility of infections to be $\alpha = \alpha_V = \alpha_1 = \alpha_2 = 1$, and therefore only modulate the relative susceptibility to disease $\epsilon$. For the initial conditions of all simulations, we take $I_P = 1 \times 10^{-9}$ and assume the remainder of the population is in the fully susceptible class. The values of the remaining parameters used in the various simulations are specified throughout the main text.

### Determination of seasonal reproduction numbers

In order to reflect observed seasonal variation in transmission rates for respiratory infections arising from related coronaviruses (*3*), influenza (*3*) and respiratory syncytial virus (*5*), we base seasonal reproduction numbers in this work on those in (*1*), which were calculated in (*3*) based on the climate of New York City. Other seasonal patterns will be possible to explore upon publication using an interactive online application. In all simulations, we modify these values to force a mean value for the basic reproduction number of $\bar{R}_0 = \langle R_0(t) \rangle = 2.3$ by multiplying the climate-derived time series $R_{0,c}(t)$ by 2.3 and dividing by its average value, i.e.

$$R_0(t) = R_{0,c}(t) \frac{2.3}{\bar{R}_{0,c}}.$$

### Modeling of nonpharmaceutical interventions (NPIs)

In all simulations, we enforce periods of NPI adoption (arising from behaviours and policies such as lock downs, mask-wearing, and social distancing) in which the transmission rate is reduced from its seasonal value described in the previous section. In particular, we assume that NPIs are adopted between weeks 8 and 47 following the pandemic onset resulting in the transmission rate being reduced to $45\%$ of its seasonal value. Between weeks 48 and 79, we assume that the transmission rate is to 30% higher than the previous time interval (reflecting an overall reduction to 45(1.3)=58.5% of the original transmission rate), due to either behavioural changes following the introduction of the vaccine or the emergence of more transmissible strains. Finally, we assume that NPIs are completely relaxed beyond week 80.

### Linking vaccination rate to inter-dose period

We consider an exponential relationship between the rate of administration of the first vaccination dose $\nu[\omega]$ and the inter-dose period $\frac{1}{\omega}$. We assume that this rate is maximized at $\nu_0$ when no second dose occurs (i.e. $\omega = 0$, an infinite inter-dose period), and that when the first and second doses are spaced by the clinically recommended inter-dose period $L_{\text{opt}}$ ($\omega_{\text{opt}} = \frac{1}{L_{\text{opt}}}$), the rate of administration of the first dose is one half of its maximum value. Thus,

$$\nu[\omega] = 2^{-L_{\text{opt}}\omega}\nu_0. \tag{S2}$$

### Calculation of the basic reproduction number $\mathcal{R}_0$

Assume that $\alpha = \alpha_V = \alpha_1 = \alpha_2 = 1$, and let $I_T = I_P + I_S + I_V + I_{S_1} + I_{S_2}$.

#### Case 1: Individuals are only vaccinated once before infection

We first assume that $x_1 = x_2 = 0$, i.e. $S_{S_1}$ and $S_{S_2}$ individuals are not vaccinated again unless they recover from infection. This corresponds to a medium-term scenario without a boosting schedule (excluding the administration of a second dose), in which it is reasonable to assume that individuals will only receive one complete vaccine regimen. In our model, this is equivalent to vaccinating $S_{S_1}$ and $S_{S_2}$ individuals without changing their immune statuses.

When there is no disease, i.e. $I_T = 0$, the governing equations reduce to

$$\frac{dS_P}{dt} = \mu - (\nu + \mu)S_P, \tag{S3a}$$

$$\frac{dV_1}{dt} = \nu S_P - (\omega + \rho_1 + \mu)V_1, \tag{S3b}$$

$$\frac{dV_2}{dt} = \omega V_1 - (\rho_2 + \mu)V_2, \tag{S3c}$$

$$\frac{dS_{S_1}}{dt} = \rho_1 V_1 - \mu S_{S_1}, \tag{S3d}$$

$$\frac{dS_{S_2}}{dt} = \rho_2 V_2 - \mu S_{S_2}. \tag{S3e}$$

Thus, with $I_T = S_S = R = 0$, it follows that the disease-free equilibrium is given by

$$S_P^{(0)} = \frac{\mu}{\mu+\nu}, \tag{S4a}$$

$$V_1^{(0)} = \frac{\mu}{\omega+\mu+\rho_1}\frac{\nu}{\mu+\nu}, \tag{S4b}$$

$$V_2^{(0)} = \frac{\mu}{\rho_2+\mu}\frac{\omega}{\omega+\mu+\rho_1}\frac{\nu}{\mu+\nu}, \tag{S4c}$$

$$S_{S_1}^{(0)} = \frac{\rho_1}{\omega+\mu+\rho_1}\frac{\nu}{\mu+\nu}, \tag{S4d}$$

$$S_{S_2}^{(0)} = \frac{\rho_2}{\rho_2+\mu}\frac{\omega}{\omega+\mu+\rho_1}\frac{\nu}{\mu+\nu}. \tag{S4e}$$

Since $I_T = I_P + I_S + I_{S_1} + I_{S_2} + I_V$, it follows from Equations (S1a)-(S1l) that

$$\frac{dI_T}{dt} = \beta I_T[S_P + \epsilon S_S + \epsilon_{V_1}V_1 + \epsilon_{V_2}V_2 + \epsilon_1 S_{S_1} + \epsilon_2 S_{S_2}] - (\gamma+\mu)I_T. \tag{S5}$$

Thus, given that $I_T'(0) < 0$ if and only if $\mathcal{R}_0 < 1$, substituting Equations (S4a)-(S4e) into Equation (S5) yields the following expression for the basic reproduction number

$$\mathcal{R}_0 = \frac{\beta}{\gamma+\mu}\left[\frac{\mu}{\mu+\nu} + \frac{\nu}{\mu+\nu}\left(\frac{\omega}{\omega+\mu+\rho_1}\left(\epsilon_{V_2}\frac{\mu}{\rho_2+\mu} + \epsilon_2\frac{\rho_2}{\rho_2+\mu}\right) + \epsilon_{V_1}\frac{\mu}{\omega+\mu+\rho_1} + \epsilon_1\frac{\rho_1}{\omega+\mu+\rho_1}\right)\right] \tag{S6}$$

Intuitively, this basic reproduction number corresponds to the fraction of individuals in each susceptible (or vaccinated) class and times their relative susceptibility (compared to a fully susceptible individual). To obtain the fraction of individuals in each susceptible/vaccinated class at the disease-free equilibrium, consider the following. First, $\frac{\mu}{\nu+\mu}$ is the fraction of individuals not vaccinated, whereas $\frac{\nu}{\nu+\mu}$ is the fraction of vaccinated individuals. This latter quantity is further split according to their immune statuses. A fraction $\frac{\omega}{\omega+\mu+\rho_1}$ of vaccinated individuals have received both doses. These individuals are further split between $V_2$ and $S_{S_2}$, i.e. a fraction $\frac{\mu}{\rho_2+\mu}$ of these individuals who have received both doses remains in $V_2$, whereas $\frac{\rho_2}{\rho_2+\mu}$ of them are in $S_{S_2}$. On the other hand, $1 - \frac{\omega}{\omega+\mu+\rho_1}$ of vaccinated individuals have not received both doses: $\frac{\mu}{\omega+\mu+\rho_1}$ of them are in $V_1$, and $\frac{\rho_1}{\omega+\mu+\rho_1}$ are in $S_{S_1}$. Thus, multiplying through gives the fraction of individuals with each respective immune status.

#### Case 2: All "susceptible" individuals are vaccinated

The previous assumption that individuals are only vaccinated once before infection is valid on a short-to medium-term timescale. However, in the longer-term, individuals will be able to be re-vaccinated as their vaccinal immunity wanes and possible boosting schedules are established. In this section, we assume instead that $S_{S_1}$ and $S_{S_2}$ individuals are vaccinated with one dose at rate $\nu$, i.e. $x_1 = x_2 = 1$, and $p_1 = 1$ and $p_2 = 0$ such that individuals in $S_{S_1}$ and $S_{S_2}$ re-enter $V_1$ and $V_2$, respectively. When there is no disease (i.e. $I_T = 0$), then

$$\frac{dS_P}{dt} = \mu - (\nu + \mu)S_P, \tag{S7a}$$
$$\frac{dV_1}{dt} = \nu(S_P + S_{S_1}) - (\omega + \rho_1 + \mu)V_1, \tag{S7b}$$
$$\frac{dV_2}{dt} = \nu S_{S_2} + \omega V_1 - (\rho_2 + \mu)V_2, \tag{S7c}$$
$$\frac{dS_{S_1}}{dt} = \rho_1 V_1 - (\mu + \nu)S_{S_1}, \tag{S7d}$$
$$\frac{dS_{S_2}}{dt} = \rho_2 V_2 - (\mu + \nu)S_{S_2}, \tag{S7e}$$

Solving these at the disease-free equilibrium gives that

$$S_P^{(0)} = \frac{\mu}{\mu + \nu}, \tag{S8a}$$
$$V_1^{(0)} = \frac{\mu}{\omega + \mu + \frac{\mu}{\nu+\mu}\rho_1}\frac{\nu}{\mu + \nu}, \tag{S8b}$$
$$V_2^{(0)} = \frac{\mu}{\frac{\mu}{\mu+\nu}\rho_2 + \mu}\frac{\omega}{\omega + \mu + \frac{\mu}{\nu+\mu}\rho_1}\frac{\nu}{\mu + \nu}, \tag{S8c}$$
$$S_{S_1}^{(0)} = \frac{\rho_1}{\omega + \mu + \frac{\mu}{\mu+\nu}\rho_1}\frac{\nu}{\mu + \nu}\frac{\mu}{\mu + \nu}, \tag{S8d}$$
$$S_{S_2}^{(0)} = \frac{\rho_2}{\frac{\mu}{\mu+\nu}\rho_2 + \mu}\frac{\omega}{\omega + \mu + \frac{\mu}{\mu+\nu}\rho_1}\frac{\nu}{\mu + \nu}\frac{\mu}{\mu + \nu}. \tag{S8e}$$

In this case, the basic reproduction number is

$$\begin{aligned}\mathcal{R}_0 = \frac{\beta}{\gamma + \mu}\Bigg[\frac{\mu}{\mu + \nu} + \frac{\nu}{\mu + \nu}\Bigg(\frac{\omega}{\omega + \mu + \frac{\mu}{\nu+\mu}\rho_1}\Bigg(\epsilon_{V_2}\frac{\mu}{\frac{\mu}{\mu+\nu}\rho_2 + \mu} + \epsilon_2\frac{\rho_2}{\frac{\mu}{\mu+\nu}\rho_2 + \mu}\frac{\mu}{\mu + \nu}\Bigg) \\ + \epsilon_{V_1}\frac{\mu}{\omega + \mu + \frac{\mu}{\nu+\mu}\rho_1} + \epsilon_1\frac{\rho_1}{\omega + \mu + \frac{\mu}{\mu+\nu}\rho_1}\frac{\mu}{\mu + \nu}\Bigg)\Bigg]\end{aligned} \tag{S9}$$

In this case, the intuition behind the formula for the basic reproduction number is as described in the previous section. However, note that the immune waning rates ($\rho_1$, $\rho_2$) in the denominators now have a coefficient $\frac{\mu}{\mu+\nu}$ (instead of 1), which is due to the re-vaccination of $S_{S_1}$ and $S_{S_2}$ individuals.

### Minimal vaccination rate for $\mathcal{R}_0 < 1$

Here we compute $\nu_{\min}$ in both scenarios examined above, i.e. when $\{1\}$ $S_{S_1}$ and $S_{S_2}$ individuals are not vaccinated and $\{2\}$ when they are vaccinated and a single dose leads to these individuals flowing back to their respective vaccinated compartments.

#### Case 1: Individuals are only vaccinated once before infection

We let $a_0 = \frac{\beta}{\gamma+\mu}$ and

$$a_1 = \epsilon_{V_1}\frac{\mu}{\omega+\mu+\rho_1} + \epsilon_{V_2}\frac{\mu}{\rho_2+\mu}\frac{\omega}{\omega+\mu+\rho_1} + \epsilon_1\frac{\rho_1}{\omega+\mu+\rho_1} + \epsilon_2\frac{\rho_2}{\rho_2+\mu}\frac{\omega}{\omega+\mu+\rho_1}, \quad \text{(S10)}$$

where it is obvious $a_1 \leq 1$ (since $\epsilon_{V_1}, \epsilon_{V_2}, \epsilon_1, \epsilon_2 \leq 1$) . Thus, $\mathcal{R}_0 = a_0\left[\frac{1}{\mu+\nu}(\mu + a_1\nu)\right]$. Since $a_1 \leq 1$, it follows that

$$\frac{d\mathcal{R}_0}{d\nu} = -\frac{a_0}{(\mu+\nu)^2}(\mu + a_1\nu) + \frac{a_0 a_1}{\mu+\nu} \leq 0. \quad \text{(S11)}$$

Thus, $\nu_{\min}$ is such that $\mathcal{R}_0 = 1$, i.e.

$$\nu_{\min} = \frac{\mu(a_0 - 1)}{1 - a_0 a_1}. \quad \text{(S12)}$$

#### Case 2: All "susceptible" individuals are vaccinated

Defining again $a_0 = \frac{\beta}{\gamma+\mu}$, we let $a = \frac{\mu}{\mu+\nu}$, i.e. $\nu = \left(\frac{1}{a} - 1\right)\mu$ so that $\mathcal{R}_0$ is now a function of $a$, i.e.

$$\begin{aligned}\mathcal{R}_0[a] = \frac{\beta}{\gamma+\mu}\Bigg(a + (1-a)\Bigg[&\epsilon_{V_1}\frac{\mu}{\omega+\mu+a\rho_1} + \epsilon_{V_2}\frac{\mu}{a\rho_2+\mu}\frac{\omega}{\omega+\mu+a\rho_1} \\ &+ \epsilon_1\frac{\rho_1}{\omega+\mu+a\rho_1}a + \epsilon_2\frac{\rho_2}{a\rho_2+\mu}\frac{\omega}{\omega+\mu+a\rho_1}a\Bigg]\Bigg).\end{aligned} \quad \text{(S13)}$$

Subsequently, we solve $\mathcal{R}_0[a] = 1$ for $\widehat{a}$ and then $\nu_{\min} = \left(\frac{1}{\widehat{a}} - 1\right)\mu$.

we can rewrite $\mathcal{R}_0[a] = 1$ as

$$f(a) = A_3a^3 + A_2a^2 + A_1a + A_0 = 0, \quad \text{(S14)}$$

where

$$A_0 = \mu(\omega+\mu)\left(\frac{1}{a_0} - \epsilon_{V_1}\frac{\mu}{\mu+\omega} - \epsilon_{V_2}\frac{\omega}{\mu+\omega}\right), \quad \text{(S15a)}$$

$$A_1 = \frac{1}{a_0}(\rho_2(\mu+\omega) + \rho_1\mu) - \mu(\omega+\mu) - \epsilon_{V_1}\mu\rho_2 - \epsilon_1\rho_1\mu - \epsilon_2\rho_2\omega + \mu(\mu\epsilon_{V_1} + \omega\epsilon_{V_2}), \quad \text{(S15b)}$$

$$A_2 = \frac{\rho_1\rho_2}{a_0} - (\mu+\omega)\rho_2 - \rho_1\mu - \epsilon_1\rho_1\rho_2 + \epsilon_1\rho_1\mu + \epsilon_2\rho_2\omega + \epsilon_{V_1}\mu\rho_2, \quad \text{(S15c)}$$

$$A_3 = -\rho_1\rho_2(1 - \epsilon_1). \quad \text{(S15d)}$$

Note that $f(a) = 0$ may have more than one feasible solution $0 < a < 1$, in which case $\nu_{\min} = \max_i\{\nu^{(i)}\}$ where $\nu^{(i)} = \left(\frac{1}{a^{(i)}} - 1\right)\mu$ for $a^{(i)}$ such that $f(a^{(i)}) = 0$. In particular, since $f(a)$ is a cubic, then $f(a) = 0$ has at most three positive roots. Thus, if $\lim_{\nu\to\infty}\mathcal{R}_0[\nu] < 1$, then this $\nu_{\min}$ is well-defined. Note that it is possible for $\mathcal{R}_0[\nu_{\min}]$ to be a local maximum ($\mathcal{R}_0'[\nu_{\min}] < 0$), in which case we still select $\nu_{\min}$.

##### $\mathcal{R}_0$ as a non-monotonic function of vaccination

In contrast to the model of (*1*), it is possible that, when all susceptible individuals are vaccinated, $\mathcal{R}_0[\nu]$ is a non-monotonic function of the vaccination rate $\nu$ (Fig. S14). This effect may occur if the following occur together: $\{1\}$ the inter-dose period is large, and $\{2\}$ the duration of complete immunity following one dose is short but the reduction in susceptibility after waning is large. Thus, these two factors may together combine to reduce the likelihood of entering $V_2$. However, as the vaccination rate $\nu$ increases, the flow of individuals changes, and it may be that more individuals are in $V_2$ and thus also in $S_{S_2}$, leading to a higher $\mathcal{R}_0$. In turn, this non-monotonic dependence of $\mathcal{R}_0$ on $\nu$ can lead to multiple solutions to $\mathcal{R}_0[\nu] = 1$ (see left panel of Fig. S14). It should be noted that conditions $\{1\}$ and $\{2\}$ together form a very pessimistic scenario, but provides an example of the possible intricacies surrounding timing of vaccine doses and corresponding immune responses.

### Vaccine refusal

In this section, we assume that $\alpha = \alpha_1 = \alpha_2 = \alpha_V = 1$, and that all susceptible individuals are vaccinated including individuals whose vaccinal immunity has waned (i.e. $x_1 = x_2 = 1$, $p_1 = 1$, $p_2 = 0$). As in (*1*), we incorporate vaccine refusal through the addition of a group of individuals with no vaccination. Thus, the model now has two interacting groups ($a$ and $b$), each with $N_i$ fraction of the population, so that $N_a + N_b = 1$. We assume that the epidemiological parameters ($\beta$, $\gamma$, $\mu$, $\epsilon$, $\delta$) are the same and that $c_{ij}$ denote the contact rate of individuals of group $i$ to $j$ relative to that of vaccine-adopters. Here, we let $I_{T,i} = I_{P,i} + I_{S,i} + I_{S_{1,i}} + I_{S_{2,i}}$. Therefore, the equations follow

$$\frac{dS_{P,a}}{dt} = \mu - \beta S_{P,a}[I_{T,a} + c_{ab}I_{T,b}] - (\nu + \mu)S_{P,a}, \tag{S16a}$$

$$\frac{dS_{S,a}}{dt} = \delta R_a - \epsilon\beta S_{S,a}[I_{T,a} + c_{ab}I_{T,b}] - (\nu + \mu)S_{S,a}, \tag{S16b}$$

$$\frac{dV_{1,a}}{dt} = \nu(S_P + cS_{S,a} + S_{S_{1,a}}) - \epsilon_{V_1}\beta V_{1,a}[I_{T,a} + c_{ab}I_{T,b}] - (\omega + \rho_1 + \mu)V_{1,a}, \tag{S16c}$$

$$\frac{dV_{2,a}}{dt} = \nu((1-c)S_{S,a} + S_{S_{2,a}}) + \omega V_{1,a} - \epsilon_{V_2}\beta V_{2,a}[I_{T,a} + c_{ab}I_{T,b}] - (\rho_2 + \mu)V_{2,a}, \tag{S16d}$$

$$\frac{dS_{S_{1,a}}}{dt} = \rho_1 V_{1,a} - \epsilon_1\beta S_{S_{1,a}}[I_{T,a} + c_{ab}I_{T,b}] - (\nu + \mu)S_{S_{1,a}}, \tag{S16e}$$

$$\frac{dS_{S_{2,a}}}{dt} = \rho_2 V_{2,a} - \epsilon_2\beta S_{S_{2,a}}[I_{T,a} + c_{ab}I_{T,b}] - (\nu + \mu)S_{S_{2,a}}, \tag{S16f}$$

$$\frac{dI_{T,a}}{dt} = \beta(S_{P,a} + \epsilon S_{P,a} + \epsilon_1 S_{S_{1,a}} + \epsilon_2 S_{S_{2,a}} + \epsilon_{V_1}V_{1,a} + \epsilon_{V_2}V_{2,a})[I_{T,a} + c_{ab}I_{T,b}] \\ \text{-}(\gamma + \mu)I_{T,a}, \tag{S16g}$$

$$\frac{dR_a}{dt} = \gamma I_{T,a} - (\delta + \mu)R_a, \tag{S16h}$$

$$\frac{dS_{P,b}}{dt} = \mu - \beta S_{P,b}[c_{ba}I_{T,a} + c_{bb}I_{T,b}] - \mu S_{P,b}, \tag{S16i}$$

$$\frac{dS_{S,b}}{dt} = \delta R_b - \epsilon\beta S_{S,b}[c_{ba}I_{T,a} + c_{bb}I_{T,b}] - \mu S_{S,b}, \tag{S16j}$$

$$\frac{dI_{T,b}}{dt} = \beta(S_{P,b} + \epsilon S_{S,b})[c_{ba}I_{T,a} + c_{bb}I_{T,b}] - (\gamma + \mu)I_{T,b}, \tag{S16k}$$

$$\frac{dR_b}{dt} = \gamma I_{T,b} - (\delta + \mu)R_b. \tag{S16l}$$

Setting $I_{T,i} = 0$, for $i = a, b$, there is a disease-free equilibrium where $S^{(0)}_{P,a} = S^{(0)}_P N_a$, $V^{(0)}_{1,a} = V^{(0)}_1 N_a$, $V^{(0)}_{2,a} = V^{(0)}_2 N_a$, $S^{(0)}_{S_{1,a}} = S^{(0)}_{S_1} N_a$, $S^{(0)}_{S_{2,a}} = S^{(0)}_{S_2} N_a$, and $S^{(0)}_{P,b} = N_b$. Using the next-generation matrix (*6, 7*), the basic reproduction number $\mathcal{R}_0$ of this model is the spectral radius $\rho$ of the matrix $K$, i.e.

$$\mathcal{R}_0 = \rho(K) = \frac{\beta}{\gamma + \mu}\left[\rho\begin{pmatrix} \mathbb{S} & c_{ab}\mathbb{S} \\ c_{ba}S^{(0)}_{P,b} & c_{bb}S^{(0)}_{P,b}\end{pmatrix}\right], \tag{S17}$$

where $\mathbb{S} = S^{(0)}_{P,a} + \epsilon_{V_1}V^{(0)}_{1,a} + \epsilon_{V_2}V^{(0)}_{2,a} + \epsilon_1 S^{(0)}_{S_{1,a}} + \epsilon_2 S^{(0)}_{S_{2,a}}$. Thus, it follows that

$$\mathcal{R}_0 = \frac{1}{2}\frac{\beta}{\gamma + \mu}\left(\sqrt{(\mathbb{S} - c_{bb}S^{(0)}_{P,b})^2 + 4c_{ab}c_{ba}\mathbb{S}S^{(0)}_{P,b}} + \mathbb{S} + c_{bb}S^{(0)}_{P,b}\right). \tag{S18}$$

We let $\mathcal{R}_{0,\infty} = \lim_{\nu\to\infty}\mathcal{R}_0$. Since $\lim_{\nu\to\infty}\mathbb{S} = \epsilon_{V_1}\frac{\mu N_a}{\omega+\mu} + \epsilon_{V_2}\frac{\omega N_a}{\omega+\mu}$, it follows that

$$\mathcal{R}_{0,\infty} = \frac{1}{2}\frac{\beta}{\gamma + \mu}\left(\sqrt{\left(\epsilon_{V_1}\frac{\mu N_a}{\omega + \mu} + \epsilon_{V_2}\frac{\omega N_a}{\omega + \mu} - c_{bb}N_b\right)^2 + 4c_{ab}c_{ba}\left(\epsilon_{V_1}\frac{\mu N_a}{\omega + \mu} + \epsilon_{V_2}\frac{\omega N_a}{\omega + \mu}\right)N_b} + \left(\epsilon_{V_1}\frac{\mu N_a}{\omega + \mu} + \epsilon_{V_2}\frac{\omega N_a}{\omega + \mu}\right) + c_{bb}N_b\right), \tag{S19}$$

where $N_a = 1 - N_b$. Then, $\mathcal{R}_{0,\infty}[N_b]$ is a function of the fraction of vaccine refusers $N_b$. We define $N_{b,\min}$ as the smallest fraction such that for all $N_b > N_{b,\min}$, then $\mathcal{R}_{0,\infty} > 1$. In other words, for all $N_b > N_{b,\min}$, there is no vaccination rate $\nu_{\min}$ as we previously defined, i.e. where any $\nu > \nu_{\min}$ leads to herd immunity ($\mathcal{R}_0 < 1$). Letting $\mathcal{S} = \epsilon_{V_1}\frac{\mu}{\omega+\mu} + \epsilon_{V_2}\frac{\omega}{\omega+\mu}$, $X = \frac{\beta}{2(\gamma+\mu)}$ and setting $\mathcal{R}_{0,\infty} < 1$ gives

$$f(N_b) = A_2 N_b^2 + A_1 N_b + A_0, \tag{S20}$$

where $f(N_b) > 0$ if and only if $\mathcal{R}_{0,\infty} < 1$. The coefficients $A_i$ are defined as

$$A_2 = (\mathcal{S} - c_{bb})^2 + [4c_{ab}c_{ba}\mathcal{S} - (\mathcal{S} + c_{bb})^2], \tag{S21a}$$

$$A_1 = \frac{2}{X}(1 - \mathcal{S}X)(\mathcal{S} - c_{bb}) - 4c_{ab}c_{ba}\mathcal{S} + 2\mathcal{S}(\mathcal{S} + c_{bb}), \tag{S21b}$$

$$A_0 = \frac{1}{X^2}(1 - 2\mathcal{S}X). \tag{S21c}$$

Thus, $f(0) = A_0 > 0$ if and only if $\mathcal{S}\frac{\beta}{\gamma+\mu} < 1$ and $f(1) = A_0 + A_1 + A_2 = \frac{1}{X^2}(1 - 2c_{bb}X) < 0$ if and only if $c_{bb}\frac{\beta}{\gamma+\mu} > 1$.

In the biologically realistic case that vaccination can lead to eradication in the absence of vaccine refusal, then $\mathcal{S}\frac{\beta}{\gamma+\mu} < 1$. Additionally, it is also biologically realistic that the disease can spread in a population made up solely of vaccine refusers, and so $c_{bb}\frac{\beta}{\gamma+\mu} > 1$. Thus, since $f$ is a quadratic function with $f(0) > 0$ and $f(1) < 0$, it has a unique root $0 < N_{b,\min} < 1$ such that for $0 \leq N_b < N_{b,\min}$, $f(N_b) > 0$ and so $\mathcal{R}_{0,\infty}[N_b] < 1$. This root $N_{b,\min}$ can be found with the quadratic formula.

### Other heterogeneities in vaccination deployment

The model in the preceding section explicitly addresses vaccine refusal. However, on a short- to medium-term, this model can also be used to more finely represent situations in which there are heterogeneities in vaccine deployment. Crudely, each group thus corresponds to those that are targeted for vaccination (such as either individuals with many contacts, or those at risk of severe disease), and those that are omitted from the initial vaccine deployment.

### Distribution of inter-dose period

The model as formulated implicitly assumes that the vaccine inter-dose period is exponentially distributed. In this section, we relax this assumption and show that our results are qualitatively similar when an Erlang distribution is considered.

To ascribe an Erlang distribution for the inter-dose period, it suffices to separate the $V_1$ compartment into two sub-compartments $V_{1_1}$ and $V_{1_2}$, with the flows to $S_{S_1}$ and $I_V$ identical and the duration in each $V_{1_i}$ exponentially distributed with mean $\frac{1}{2\omega}$. Thus, the (S1f) to (S1i) equations become

$$\begin{aligned}\frac{dV_{1_1}}{dt} &= s_{\text{vax}}\nu S_P + cs_{\text{vax}}\nu S_S + x_1 p_1 s_{\text{vax}}\nu S_{S_1} + x_2 p_2 s_{\text{vax}}\nu S_{S_2} \\ &\quad - \epsilon_{V_1}\beta V_{1_1}[I_P + \alpha I_S + \alpha_V I_V + \alpha_1 I_{S_1} + \alpha_2 I_{S_2}] - (2\omega + \rho_1 + \mu)V_{1_1},\end{aligned} \tag{S22a}$$

$$\frac{dV_{1_2}}{dt} = 2\omega V_{1_1} - \epsilon_{V_1}\beta V_{1_2}[I_P + \alpha I_S + \alpha_V I_V + \alpha_1 I_{S_1} + \alpha_2 I_{S_2}] - (2\omega + \rho_1 + \mu)V_{1_2}, \tag{S22b}$$

$$\begin{aligned}\frac{dV_2}{dt} &= (1-c)s_{\text{vax}}\nu S_S + x_1(1-p_1)s_{\text{vax}}\nu S_{S_1} + x_2(1-p_2)s_{\text{vax}}\nu S_{S_2} + 2\omega V_{1_2} \\ &\quad - \epsilon_{V_2}\beta V_2[I_P + \alpha I_S + \alpha_V I_V + \alpha_1 I_{S_1} + \alpha_2 I_{S_2}] - (\rho_2 + \mu)V_2,\end{aligned} \tag{S22c}$$

$$\frac{dI_V}{dt} = \beta[\epsilon_{V_1}(V_{1_1} + V_{1_2}) + \epsilon_{V_2}V_2][I_P + \alpha I_S + \alpha_V I_V + \alpha_1 I_{S_1} + \alpha_2 I_{S_2}] - (\gamma + \mu)I_V, \tag{S22d}$$

$$\frac{dS_{S_1}}{dt} = \rho_1(V_{1_1} + V_{1_2}) - \epsilon_1\beta S_{S_1}[I_P + \alpha I_S + \alpha_V I_V + \alpha_1 I_{S_1} + \alpha_2 I_{S_2}] - (s_{\text{vax}}x_1\nu + \mu)S_{S_1}, \tag{S22e}$$

In Figure S9 we present analogous scenarios as in Figure 2 of the main text for the short- and medium-term dynamics (i.e. $x_1 = x_2 = 0$) with an Erlang-distributed inter-dose period. We find that the temporal dynamics are qualitatively very similar.

### Additional scenarios

#### Different vaccination rates

In this section, we examine the impact of different first-dose administration rates on medium-term outcomes. In order to provide a baseline for the medium-term dynamics, we begin by plotting scenarios where vaccines are not deployed (Fig. S1). Next, due to the possibility that certain countries/regions may have limited access to vaccines, we model scenarios where the first-dose administration rate is maximally $0.1\%$ per week (Fig. S2). Unsurprisingly, such limited vaccine deployment efforts lead to marginal differences between dosing regimes. Both scenarios highlight the substantial benefits that the sufficiently rapid deployment of even an imperfect vaccine can have.

In Figure S3, we examine a maximum first-dose administration rate of $1\%$ (instead of $2\%$ in Figure 2). In contrast to Figure 2, the first epidemic peak post-vaccination initiation is very moderately affected by variations in dosing regimes. However, in the case of less robust natural and vaccinal immunity (Figure S3A), and a poorer immune response following one vaccine dose (Figure S3A top section), the longer term burden increases if second doses are delayed (or not administered). If vaccinal immunity following one dose is robust, then one-dose strategies lead to lower burden.

Certain countries will likely be able to vaccinate their populations quickly, and thus have a high first-dose administration rate per week. In Figure S4, we examine the short- and medium-term dynamics with a first-dose administration rate of $5\%$ per week. In the case of poorer natural and vaccinal immunity (Figure S4A), implementing a one-dose strategy when the immune response elicited by a single dose is shorter and weaker than that from two doses results in larger and delayed subsequent epidemics ((compare the leftmost and rightmost columns of the top section of Figure S4A). On the other hand, if one vaccine dose provides more robust protection (Figure S4A bottom section), then the model predicts that delaying the second dose may result in a larger first epidemic peak after the start of vaccination, although this peak may be later and fewer peaks may arise relative to the case where the recommended two-dose strategy is followed. If natural and vaccinal immunity are overall more robust (Figure S4B) and a first vaccine dose is poor (Figure S4B top section), then some intermediate delay of the second dose (larger than the recommended timing) can lead to substantially reduced infections compared to a one-dose strategy, and the recommended two-dose strategy immediately after vaccine onset (compare middle plots of the top section of Figure S4B with the leftmost and rightmost panels). If one-dose immunity is robust, then any delay leads to fewer cases than the recommended two dose strategy (Figure S4B bottom section), although case numbers are low for all dosing regimes. In all of these scenarios (assuming a a maximal $5\%$ per week rate of first-dose administration), the accumulation of $S_{S_1}$ individuals qualitatively follows our results from Figure 2 of the main text.

#### Ramp-up of vaccination

In this section, we examine highly simplified scenarios for increases in vaccine deployment.

##### Increase in vaccination administration does not affect dosage policy

The first example we consider is where the maximal rate of first-dose administration increases, but the vaccine dosing regime does not change (Figure S10). In particular, we examine a scenario where the maximal first-dose administration is initially $1\%$, but that this value then switches to $3\%$ after week 60.

##### Increase in vaccination and a shift to optimally-timed two-dose policy

As vaccine deployment increases, it is also possible that changes in vaccine dosing regimes may occur. To account for this, we examine scenarios where the maximal rate of administration of the first-dose is constant for all times, but the dosing regime is allowed to change "at no cost". In other words, all depicted vaccine dosing regimes switch to the recommended two-dose regime after a certain period, with the rate of administration of the first dose corresponding to its maximal value (i.e. the rate for a one-dose policy). As examples: if the initial dosing regime is a two-dose strategy with the recommended inter-dose spacing, then its rate of administration of the first dose doubles. On the other hand, if the initial dosing regime is a exclusively one-dose policy, then the rate administration of the first dose does not change under the new vaccine regime, but second doses are now administered using the recommended inter-dose period. These scenarios are depicted in Figures S11 and S12 for switches in dosing regimes after weeks 60 and 80, respectively.

#### Dosing strategies for vaccine deployment in populations with different prior attack rates

##### Larger initial attack rate

In the main text, we have assumed that the initial conditions at vaccination crudely match those of a North American or European city that implemented NPIs to restrict disease transmission before the initiation of vaccination. However, certain cities across the world have had large fractions of their populations already infected. (e.g. Manaus, Brazil (*8*)). In Fig. S5, we explore the dynamical impact of vaccination in a population that has largely been infected before the start of vaccination. To do so, all parameters are the same as in Figure 2 of the main text, with the exception of the NPI scenario. Here, less restrictive NPIs are adopted between weeks $11$ and $49$ inclusive only, with the transmission rate reduced to $58.5\%$ of its seasonal value during this period. As expected, epidemiological differences due to different dosing strategies are more modest both in the short- and medium-term.

##### Successfully suppressed disease spread before the onset of vaccination

Additionally, certain countries have successfully reduced transmission and experienced more contained early epidemics (e.g. New Zealand (*9*), Taiwan (*10*), Australia (*11*)).This reduced number of infections is associated with the population remaining almost completely susceptible at the time of vaccine onset, which we explore in Figure S6. All parameters correspond to those in Figure 2 of the main text with the exception of the NPI scenario. Here, we assume that strict NPIs are introduced between the onset of the pandemic and week 47 (inclusive), resulting in the transmission rate being reduced to $40\%$ of its seasonal value. Subsequently, and coinciding with vaccine onset, we assume that less strict NPIs are adopted between weeks 48 and 103, inclusive, during which the transmission rate is reduced to $58.5\%$ of its seasonal value. Beyond this we assume that NPIs are completely relaxed. Intuitively, a two-dose regime according to the recommended schedule is favourable if one-dose vaccinal immunity is less robust than that elicited after two doses, whereas delaying second doses leads to less longer-term burden if one-dose immunity is robust.

#### The timing of vaccine deployment

In Figures S7 and S8, we examine the impact of initiating vaccination after weeks 52 and 65, respectively (compared to 48 weeks in Figure 2 of the main text). A slight delay in vaccination leads to a reduction in the impact of the choice of dosing regime on the size of the first epidemic peak after vaccine onset (Figure S7), which is reminiscent of what was observed for lower rates of administration of the first vaccine dose (Figures S2 and S3). Otherwise, even with a small delay in the onset of vaccination, the general effects of vaccine dosage regimes on the medium-term dynamics are similar to those depicted in Figure 2.

Delaying vaccination until after week 65 leads to more modest differences between cases and burden across different dosing regimes, although the timing and size of future epidemic peaks are still shaped by the nature of natural and vaccinal immunity and the relative robustness of immunity elicited after the first vaccine dose. Furthermore, the fraction of susceptible individuals whose one-dose vaccinal immunity has waned is highest for dosing regimes that emphasize the first dose and delay (or omit) the second.

We note that these results are dependent on the number of infections (i.e. the point in the dynamical cycle) when vaccines are introduced, highlighting the critical interplay between the force of infection and population immunity in determining epidemiological trajectories.

### Caveats and future directions

There are numerous caveats to our work, and each gives rise to important areas for future research. Below, we provide a complete list of these caveats, which are summarized in the main text.

- Our model (and that of (*1*)) assumes exponentially-distributed immune durations. With more data, appropriate distributions of immunity could be parameterized and in turn used in our model for quantitative predictions. Additionally, we have assumed that the characteristics of infections after the waning of natural or vaccinal immunity do not change as the number of re-infections increase, but this could be further refined through numerous additional compartments in the underlying model.
- We have also assumed a very simple model for evolutionary potential, where infections after the waning of immunity each contribute to the relative net adaptation rate. In particular we ignore onward transmission of escape variants; this is an important area to explore. Coupling our immuno-epidemiological model to more accurate evolutionary models (*12, 13*), and perhaps to sequence analyses, could give more accurate quantitative evolutionary predictions. However, modeling these cross-scale dynamics remains a formidable problem (*14*).

- Our model of NPIs simply assumes constant reductions of transmission for certain time intervals, and does not contain complex temporal effects. However, we endeavour to study qualitative immuno-epidemiological dynamics in the medium-term; for accurate quantitative predictions for a specific region, our model could be fit with finely resolved NPI data. Additionally, the periods and strengths of NPIs will be able to be varied upon publication using an online interactive application.

- Numerous heterogeneities in SARS-CoV-2 transmission exist (*15–17*), and heterogeneities may impact the attainment of herd immunity (*18–20*). However, motivated by previous work that found that the underlying immuno-epidemiological model was qualitatively robust to heterogeneities (*1*), we omit most of these in our model. However, to model vaccine refusal, we do examine an extension of our model with two interacting groups in the Supplementary Materials. Additionally, we also discuss how the model for vaccine refusal can crudely represent heterogeneities in vaccine deployment. Incorporating various heterogeneities in our framework is thus an important area for future quantitative predictions.

- In our comparisons of vaccine dosing regimes, we have assumed, based on the lack of available data, that the inter-dose period is decoupled from the immune response elicited after the second dose. However, the timing of subsequent doses for other multi-dose vaccines can affect subsequent immune responses, perhaps beneficially (e.g. (*21*)). Thus, as more data become available, the scenarios we investigate could be refined by including a relationship between the robustness of immune responses following two doses and the average inter-dose period in the immuno-epidemiological model. We also do not account for the complexities of repeat dosing with different vaccines, which is an important area for future theoretical and empirical research.

- We have also assumed a constant rate of administration of the first vaccine dose, although we have investigated the impacts and implications of increasing vaccination capacity in Figures S10, S11, and S12. As vaccines are distributed, more refined vaccine deployment rates will become available and could be incorporated into our model.

- Lastly, we have assumed that vaccination occurs randomly in the population (e.g. previously-infected individuals are equally likely to receive vaccination), but it is possible that, based on their current immunological/epidemiological status, individuals may choose to obtain vaccination differently. Thus, the rate of administration of the first vaccine dose could change over time if more/fewer susceptible individuals choose to obtain a vaccine compared to individuals with current infections or immunity.

### Supplementary figures

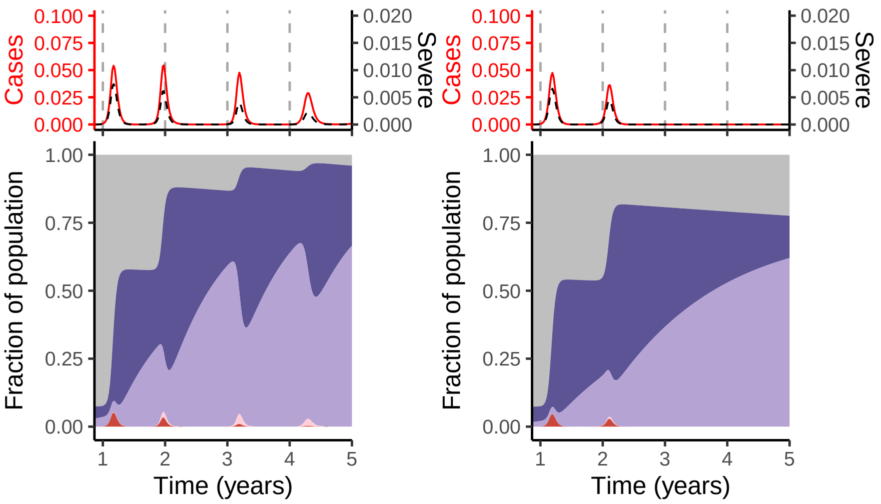

**Figure S1:** Immuno-epidemiological trajectories in the absence of vaccination, with all other parameter values and details identical to those in Figure 2 of the main text. The left panel depict trajectories in the presence of poor immunity (i.e. Figure 2A), whereas the right panel corresponds to Figure 2B and examines the absence of vaccination with robust immune responses after infection.

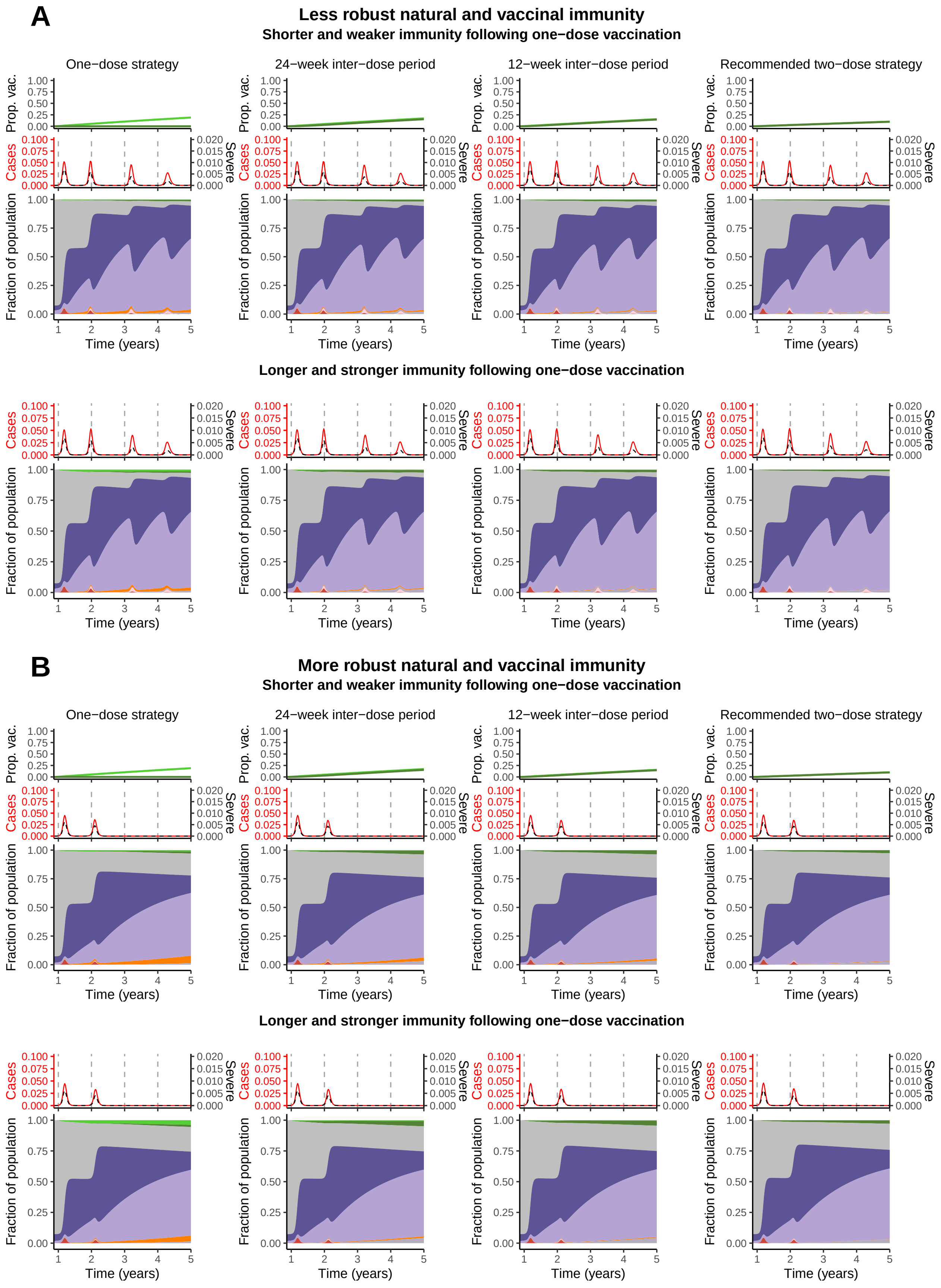

**Figure S2:** Immuno-epidemiological trajectories for a maximal rate of administrate of the first vaccine dose of $\nu_0 = 0.1\%$ per week, with all other parameter values and details identical to those in Figure 2 of the main text.

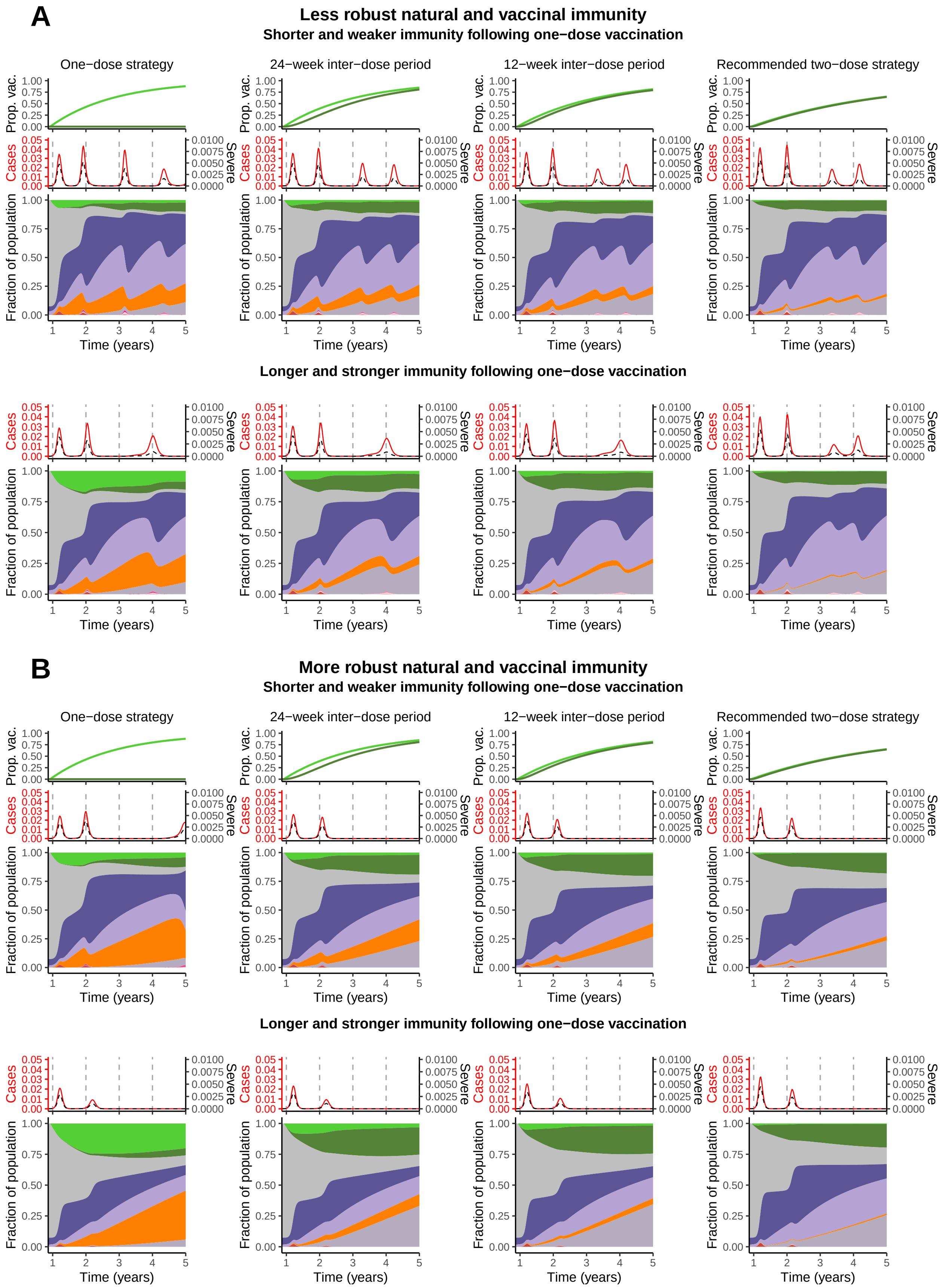

**Figure S3:** Immuno-epidemiological trajectories for a maximal rate of administrate of the first vaccine dose of $\nu_0 = 1\%$ per week, with all other parameter values and details identical to those in Figure 2 of the main text.

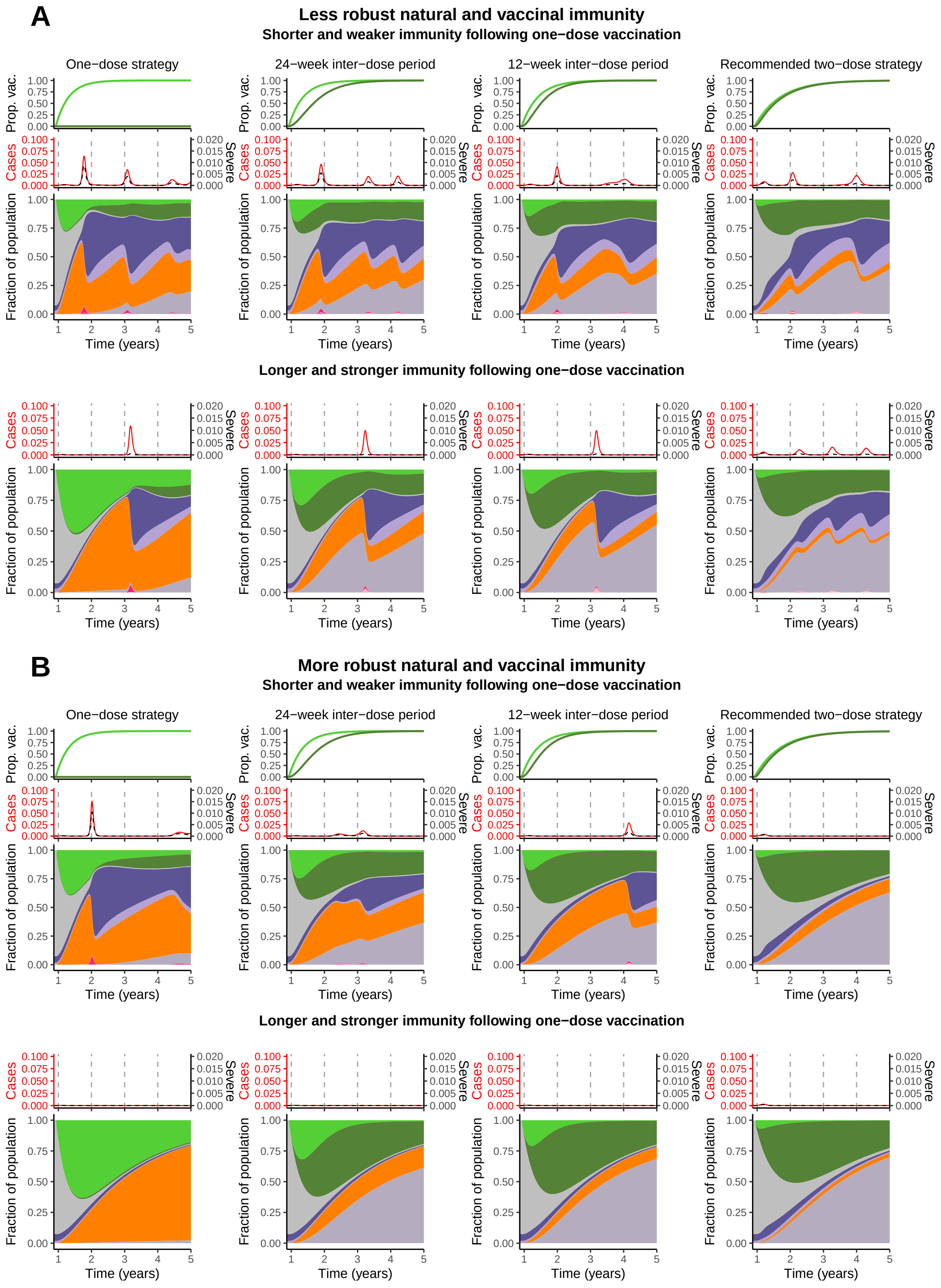

**Figure S4:** Immuno-epidemiological trajectories for a maximal rate of administrate of the first vaccine dose of $\nu_0 = 5\%$ per week, with all other parameter values and details identical to those in Figure 2 of the main text.

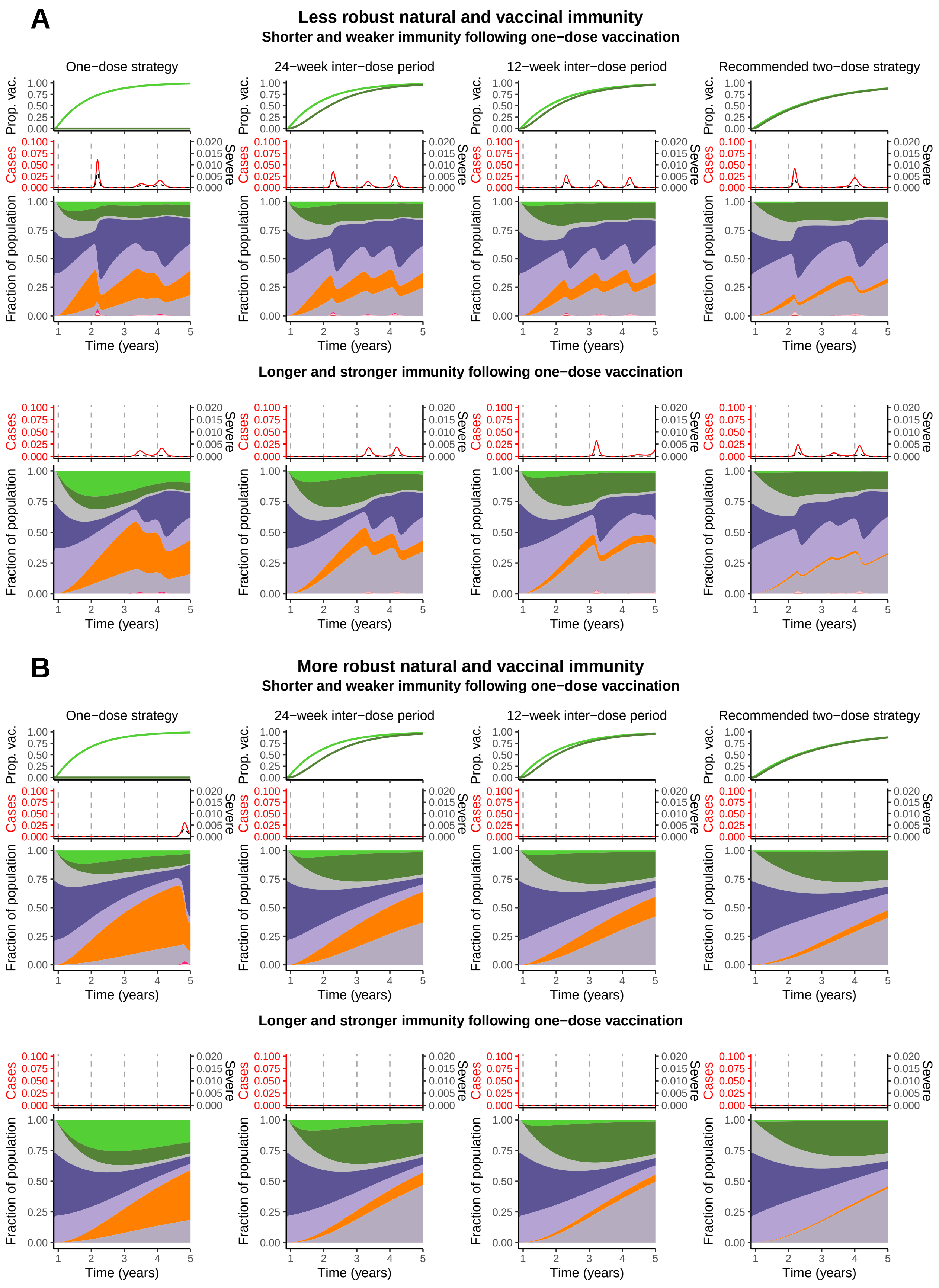

**Figure S5:** Immuno-epidemiological trajectories corresponding to a scenario in which vaccination is initiated after a large fraction of the population has been previously infected. Parameters are identical to those in Figure 2 of the main text with the exception of the NPI adoption strategy (see Supplementary Materials for details).

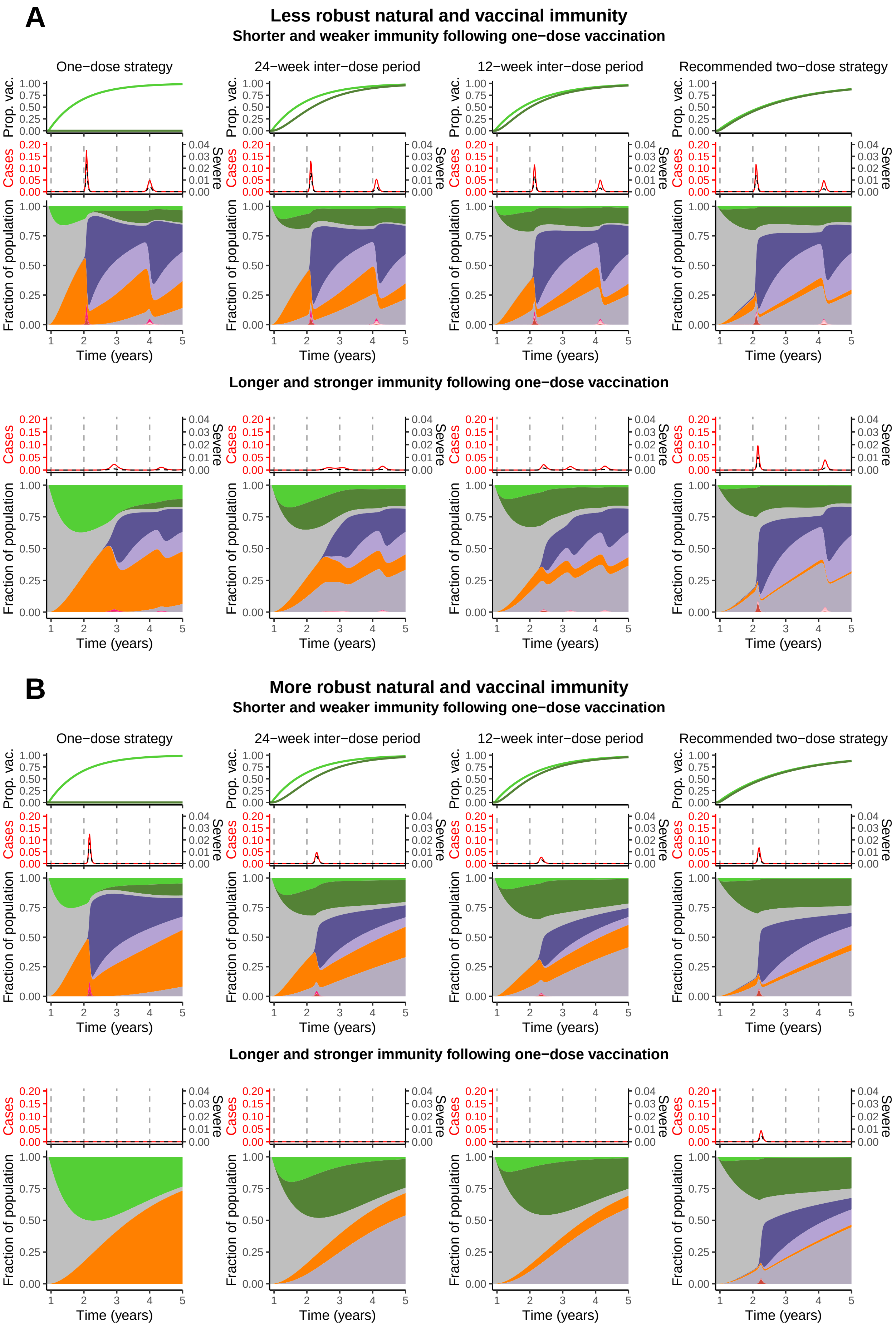

**Figure S6:** Immuno-epidemiological trajectories corresponding to a scenario in which vaccination is initiated in a population with almost no community immunity. Parameters are identical to those in Figure 2 of the main text with the exception of the NPI adoption strategy (see Supplementary Materials for details).

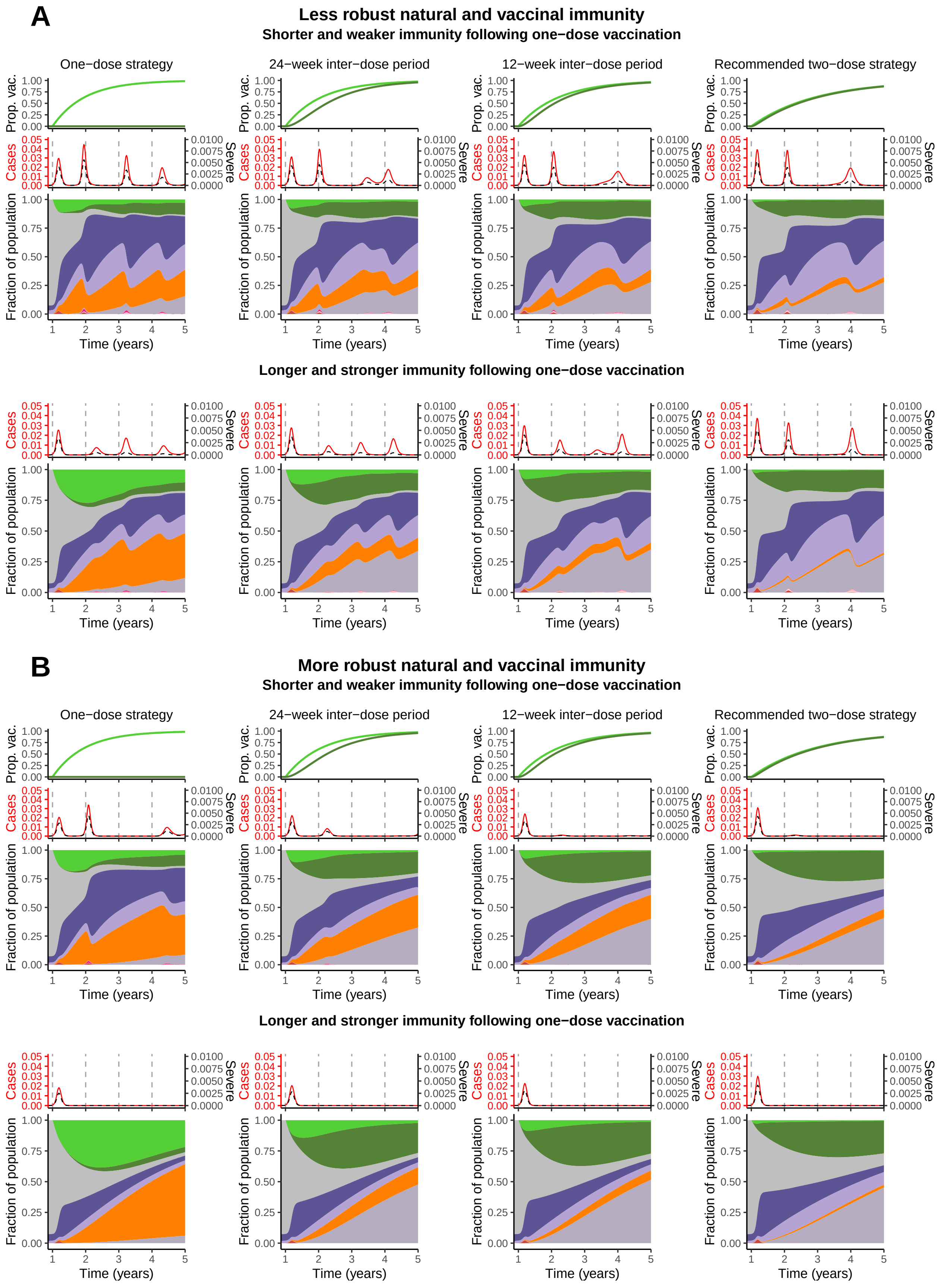

**Figure S7:** Immuno-epidemiological trajectories when vaccination is delayed until after week 52, with all other parameter values and details identical to those in Figure 2 of the main text.

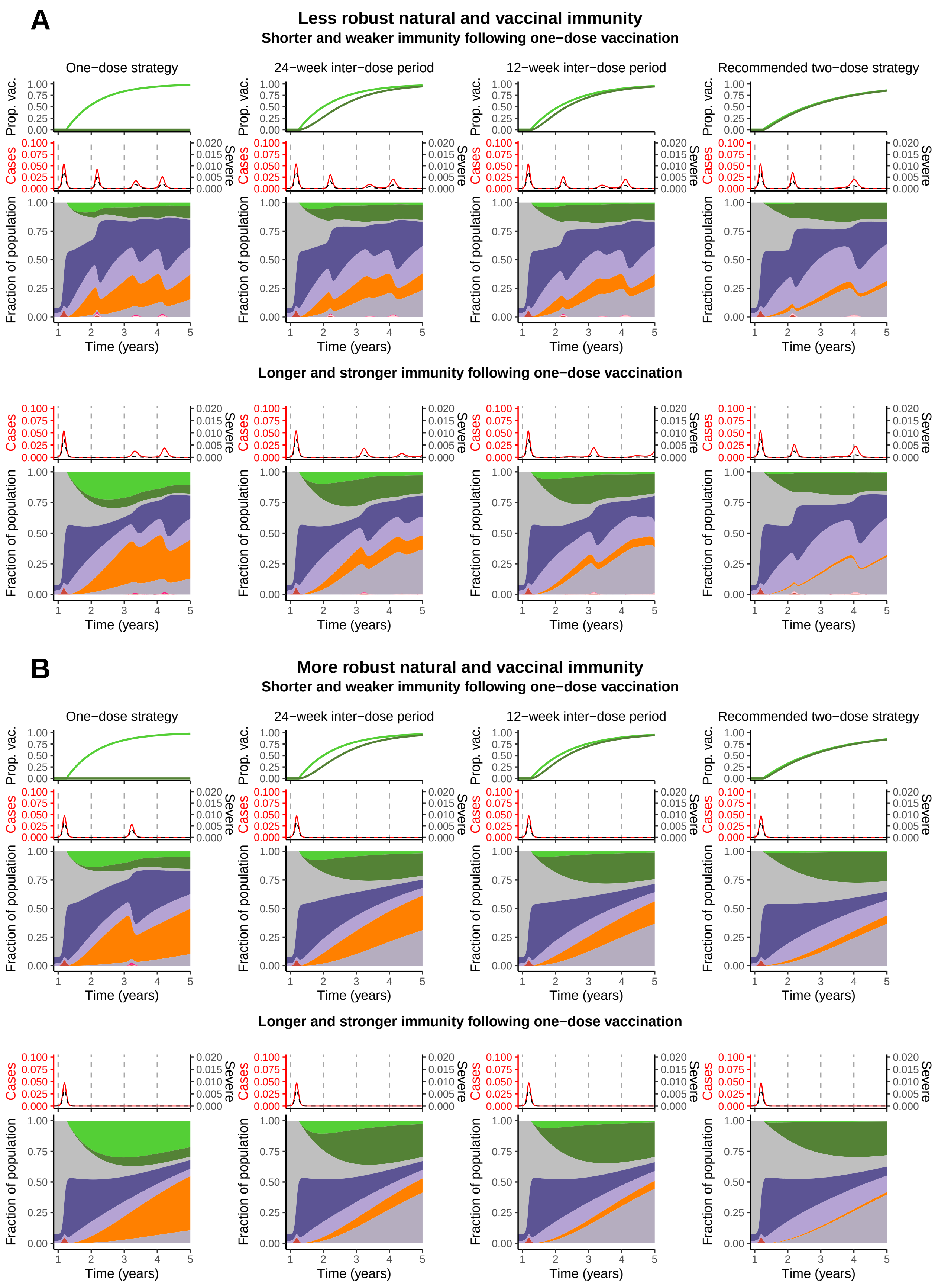

**Figure S8:** Immuno-epidemiological trajectories when vaccination is delayed until after week 65, with all other parameter values and details identical to those in Figure 2 of the main text.

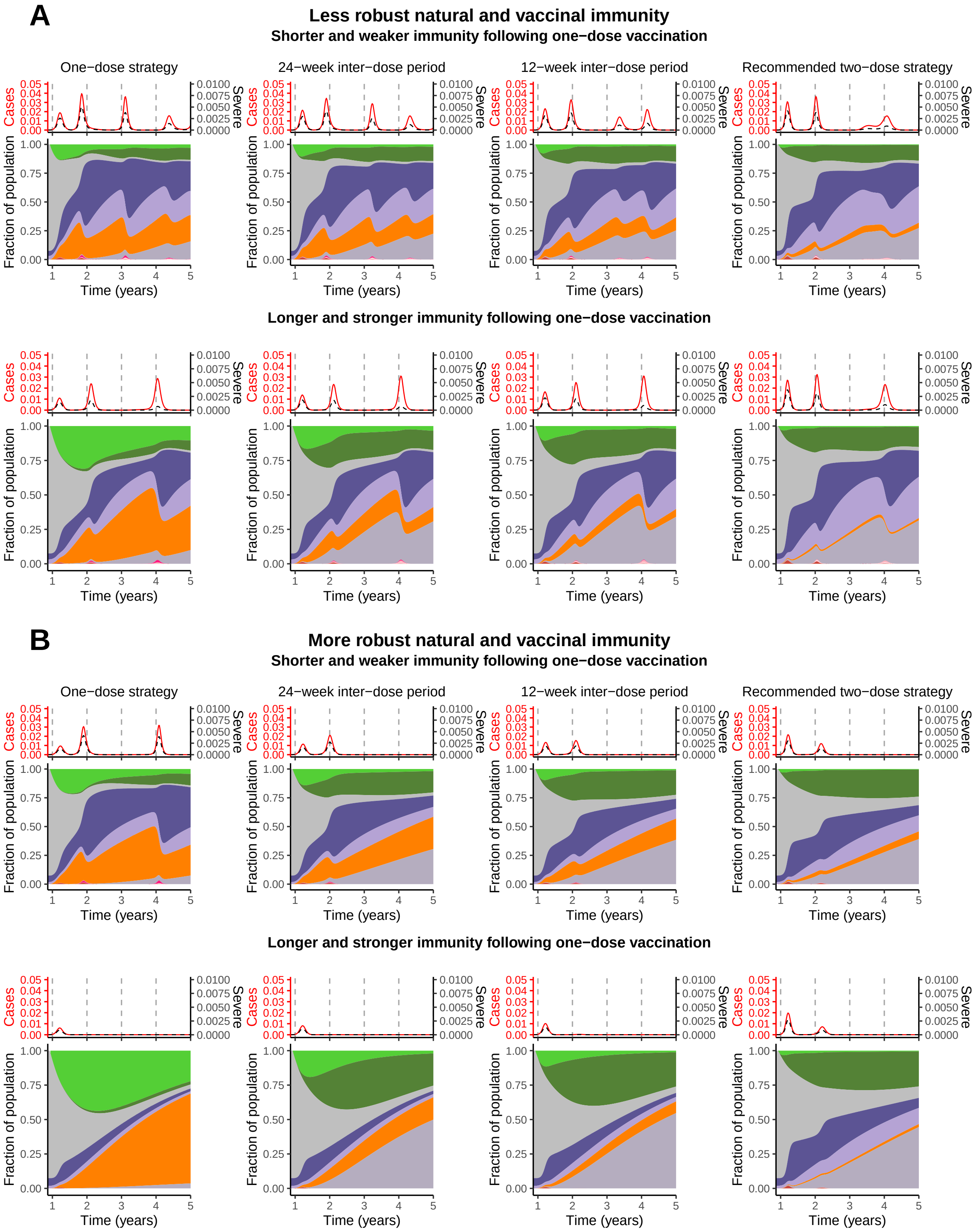

**Figure S9:** Immuno-epidemiological trajectories when an Erlang-distributed inter-dose period is considered (see Supplementary Materials). All other parameter values and details are identical to those in Figure 2 of the main text.

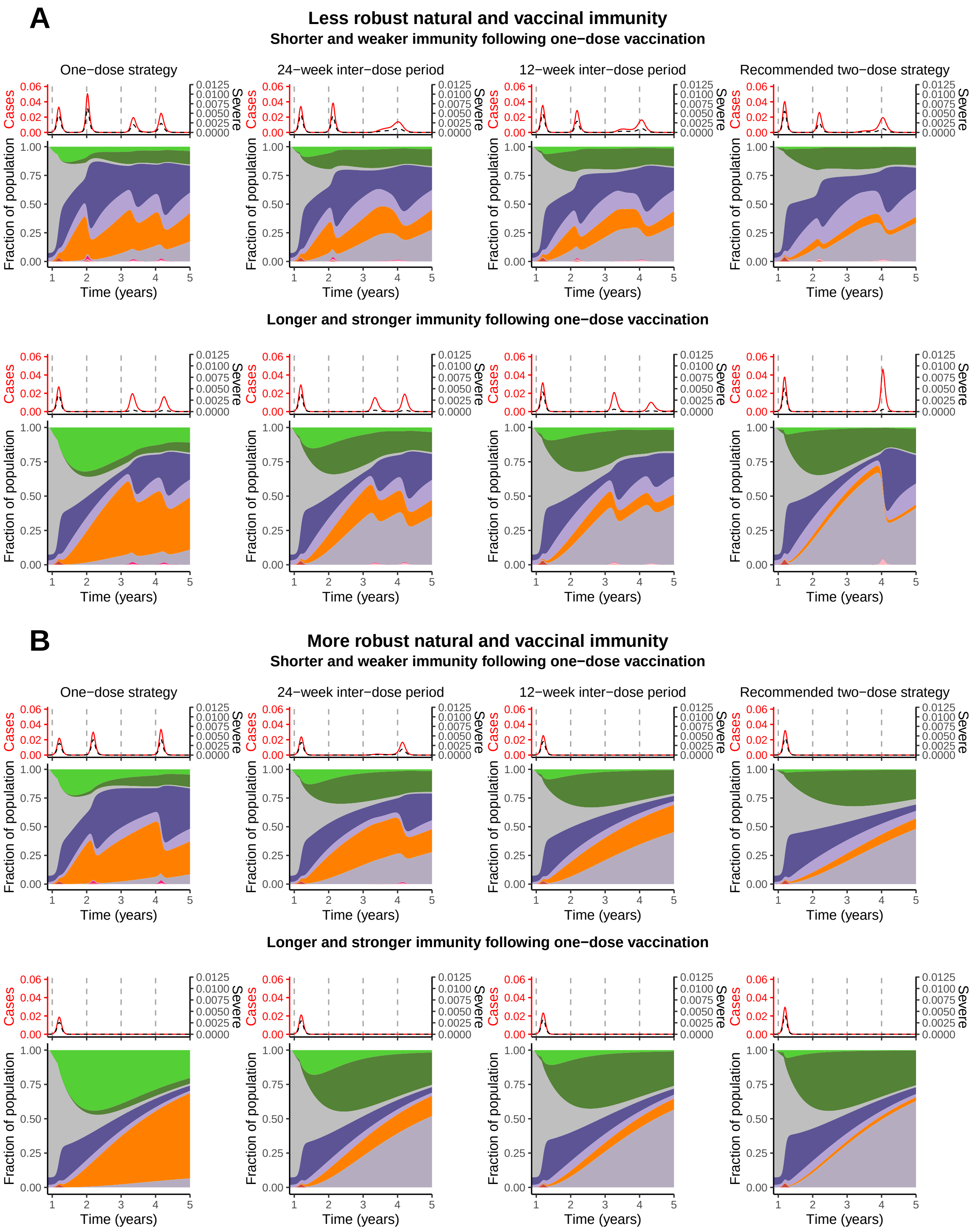

**Figure S10:** Immuno-epidemiological trajectories when the rate of administration of the first vaccine dose is increased from $\nu_0 = 1\%$ per week to $\nu_0 = 3\%$ per week after week 60. All other parameter values and details are identical to those in Figure 2 of the main text

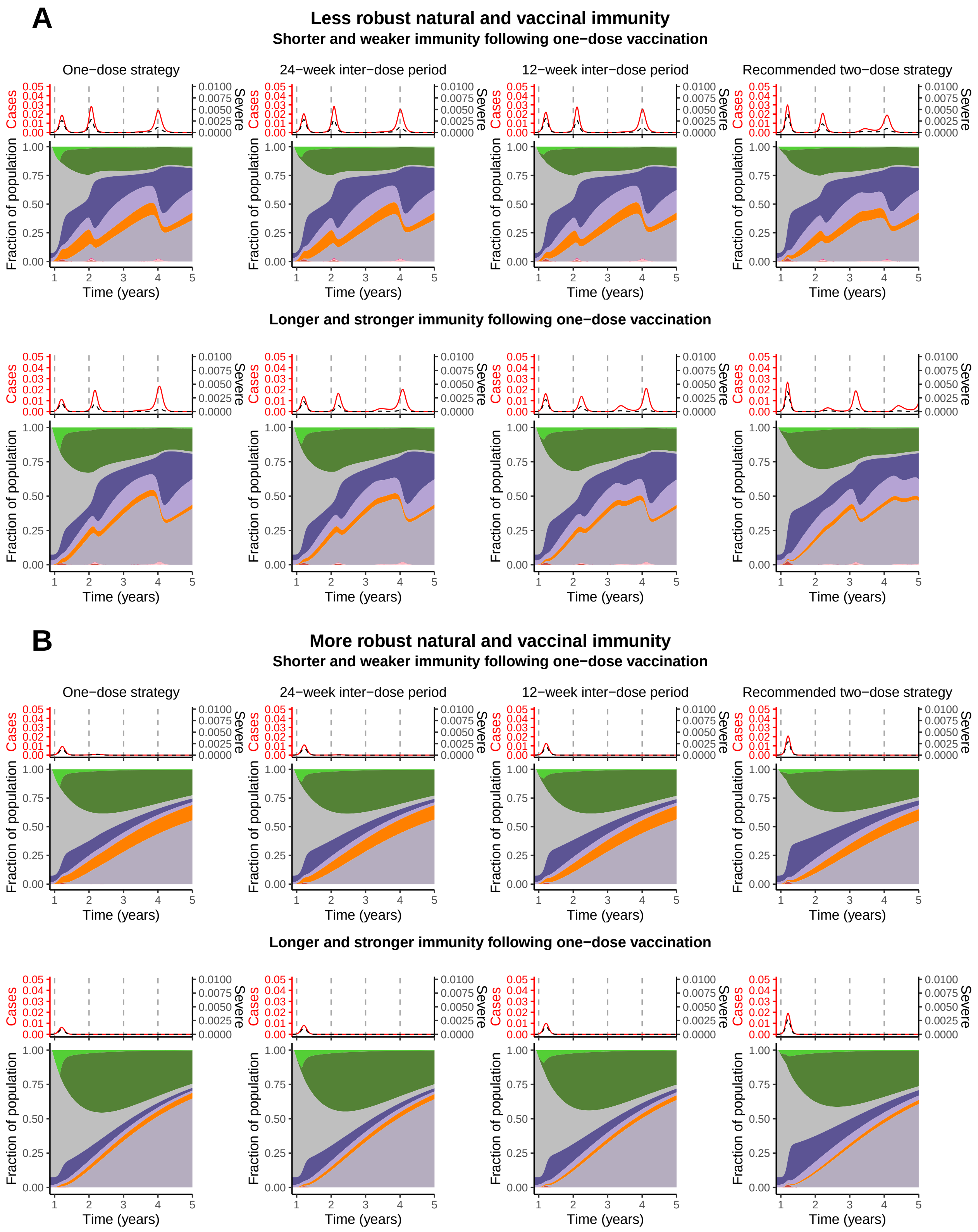

**Figure S11:** Immuno-epidemiological trajectories when the vaccine regime is changed to a two dose policy with recommended spacing after week 60, with the rate of administration of the first vaccine dose $\nu_0$ unchanged. All other parameter values and details are identical to those in Figure 2 of the main text.

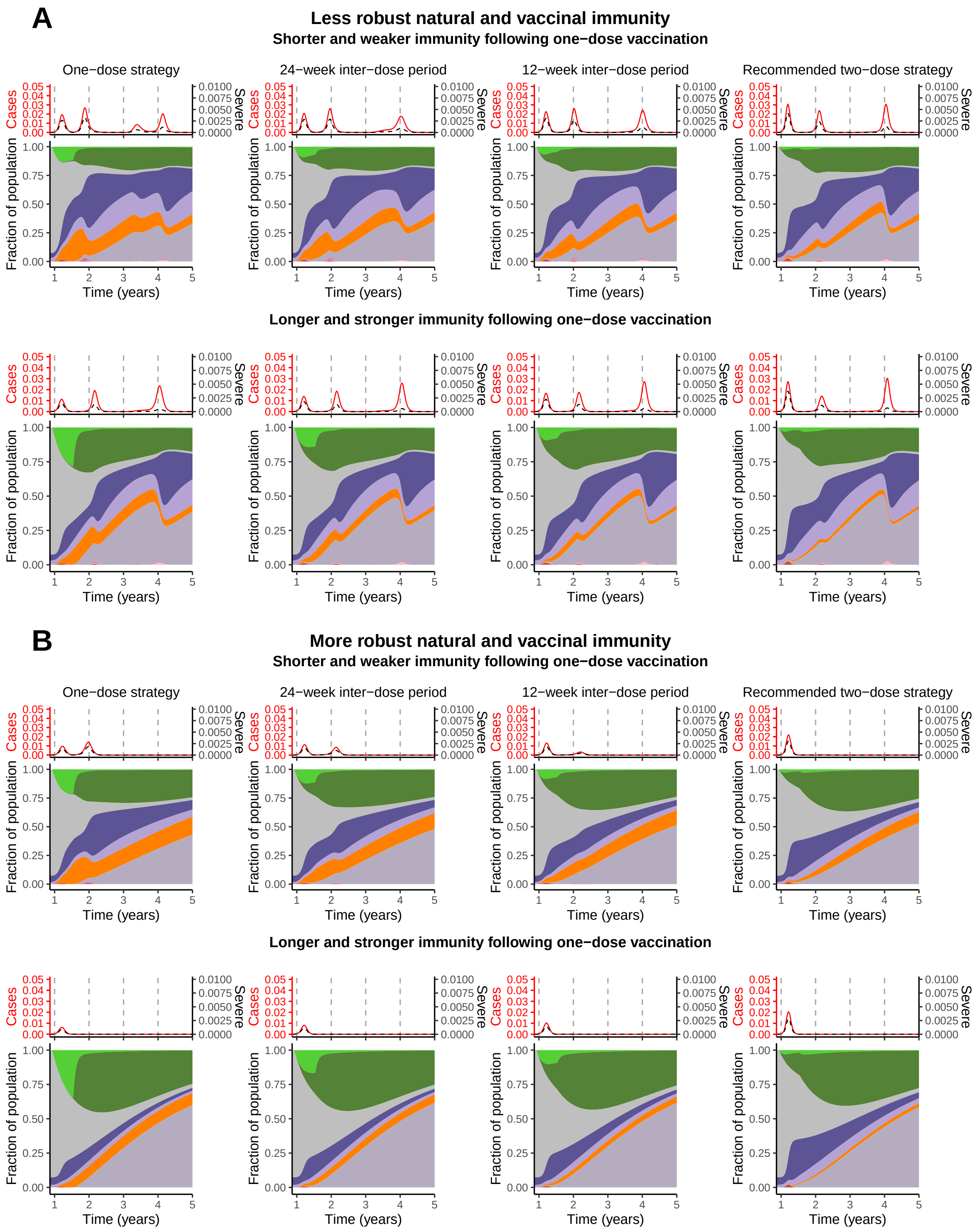

**Figure S12:** Immuno-epidemiological trajectories when the vaccine regime is changed to a two dose policy with recommended spacing after week 80, with the rate of administration of the first vaccine dose $\nu_0$ unchanged. All other parameter values and details are identical to those in Figure 2 of the main text.

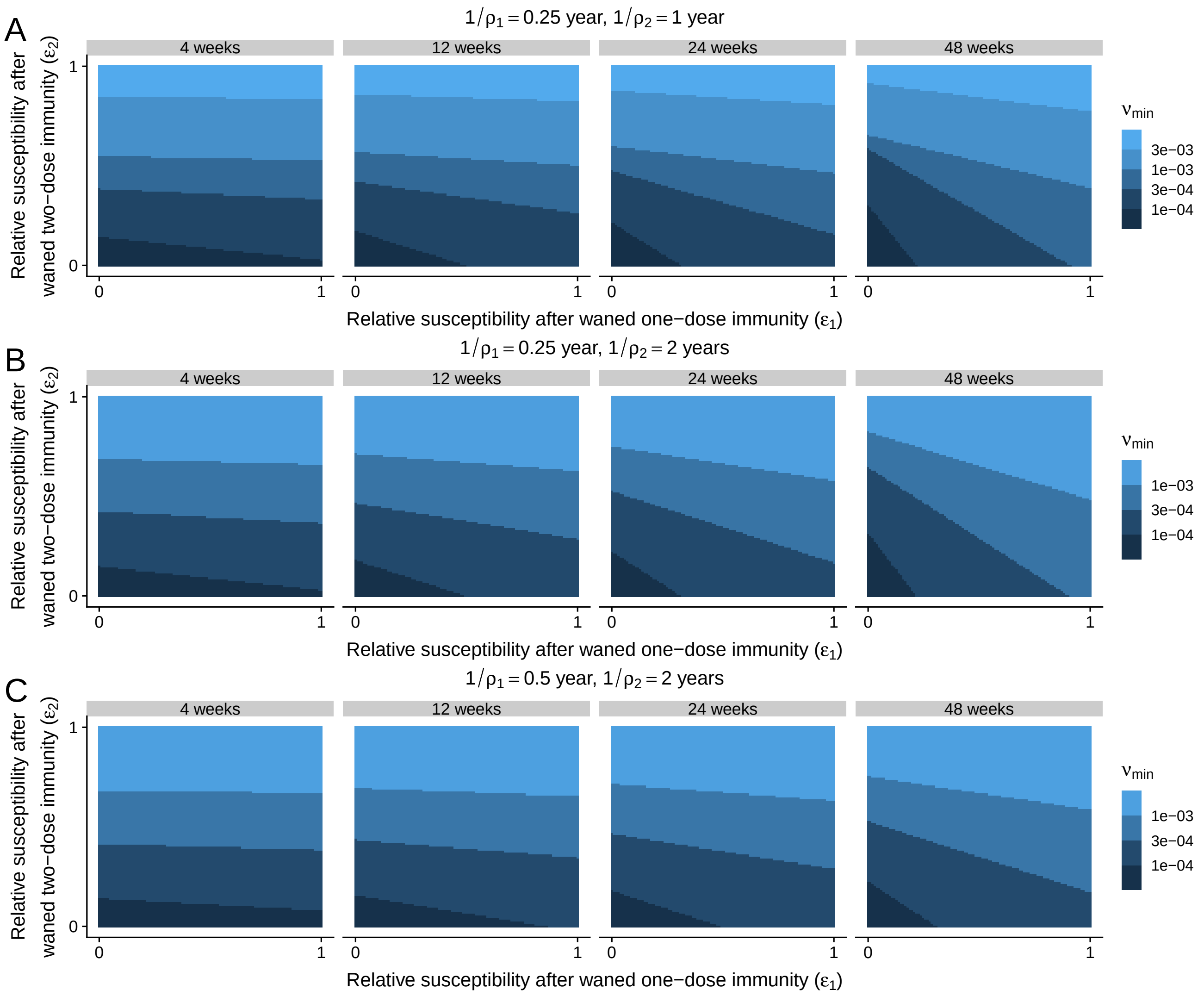

**Figure S13:** Minimum daily rate of administration of the first dose $\nu_{\text{min}}$ such that for any higher rate, $\mathcal{R}_0 < 1$. Parameter values other than $\rho_1$ and $\rho_2$ are as in Figure 3B.

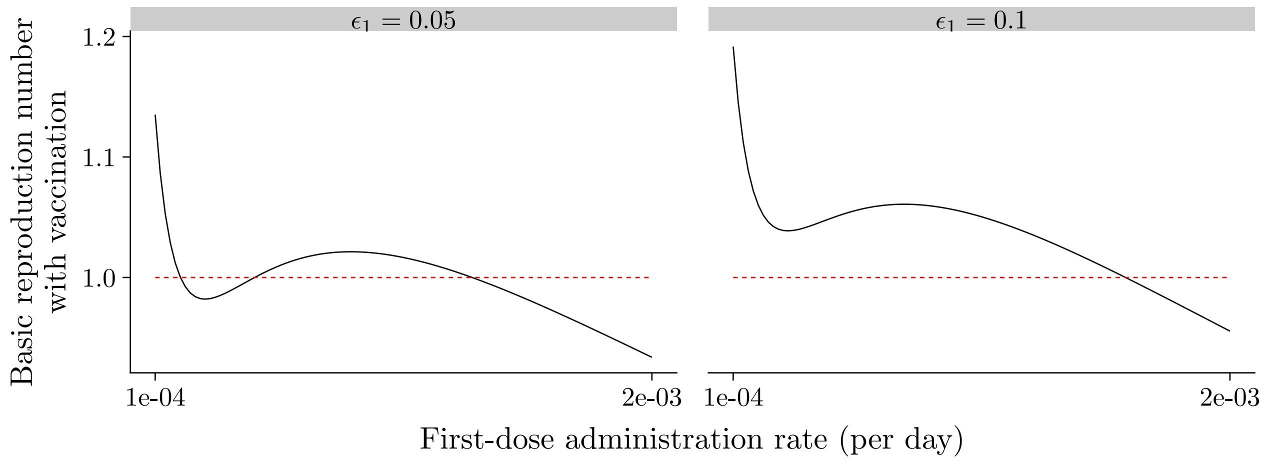

**Figure S14:** Pessimistic scenarios in which the basic reproduction number $\mathcal{R}_0$ is a non-monotonic function of the rate of administration of the first vaccine dose. Parameter values: $\omega = \frac{1}{48(7)}$ per day, $\rho_1 = \frac{1}{0.1(365)}$ per day, $\rho_2 = \frac{1}{365}$ per day, $\epsilon_2 = 0.8$, $\epsilon_{V_1} = 0.1$, $\epsilon_{V_2} = 0.05$, $\beta = \frac{2.3}{5}$ per day, $\gamma = \frac{1}{5}$ per day, and $\mu = \frac{1}{50(365)}$.

8. Lewis F. Buss, Carlos A. Prete, Claudia M. M. Abrahim, Alfredo Mendrone, Tassila Salomon, Cesar de Almeida-Neto, Rafael F. O. França, Maria C. Belotti, Maria P. S. S. Carvalho, Allyson G.

Costa, Myuki A. E. Crispim, Suzete C. Ferreira, Nelson A. Fraiji, Susie Gurzenda, Charles Whittaker, Leonardo T. Kamaura, Pedro L. Takecian, Pedro da Silva Peixoto, Marcio K. Oikawa, Anna S. Nishiya, Vanderson Rocha, Nanci A. Salles, Andreza Aruska de Souza Santos, Martirene A. da Silva, Brian Custer, Kris V. Parag, Manoel Barral-Netto, Moritz U. G. Kraemer, Rafael H. M. Pereira, Oliver G. Pybus, Michael P. Busch, Márcia C. Castro, Christopher Dye, Vítor H. Nascimento, Nuno R. Faria, and Ester C. Sabino. Three-quarters attack rate of sars-cov-2 in the brazilian amazon during a largely unmitigated epidemic. *Science*, 371(6526):288–292, 2021.

9. Michael G. Baker, Nick Wilson, and Andrew Anglemyer. Successful elimination of covid-19 transmission in new zealand. *New England Journal of Medicine*, 383(8):e56, 2020.

10. Dr Jennifer Summers, Dr Hao-Yuan Cheng, Professor Hsien-Ho Lin, Dr Lucy Telfar Barnard, Dr Amanda Kvalsvig, Professor Nick Wilson, and Professor Michael G Baker. Potential lessons from the taiwan and new zealand health responses to the covid-19 pandemic. *The Lancet Regional Health –Western Pacific*, 4, 2021/01/16 2020.

11. COVID-19 National Incident Room Surveillance Team. Epidemiology Report 31: Fortnightly reporting period ending 6 December 2020. *Commun Dis Intell*, 44, 2020. DOI:doi:10.33321/cdi.2020.44.92.

12. Neil M. Ferguson, Alison P. Galvani, and Robin M. Bush. Ecological and immunological determinants of influenza evolution. *Nature*, 422(6930):428–433, 2003.

13. Katia Koelle, Sarah Cobey, Bryan Grenfell, and Mercedes Pascual. Epochal evolution shapes the phylodynamics of interpandemic influenza a (h3n2) in humans. *Science*, 314(5807):1898–1903, 2006.

14. Chadi M Saad-Roy, Adrian B McDermott, and Bryan T Grenfell. Dynamic Perspectives on the Search for a Universal Influenza Vaccine. *The Journal of Infectious Diseases*, 219(Supplement˙1):S46–S56, 04 2019.

15. Jennie S. Lavine, Ottar N. Bjornstad, and Rustom Antia. Immunological characteristics govern the transition of covid-19 to endemicity. *Science*, 2021.

16. Kaiyuan Sun, Wei Wang, Lidong Gao, Yan Wang, Kaiwei Luo, Lingshuang Ren, Zhifei Zhan, Xinghui Chen, Shanlu Zhao, Yiwei Huang, Qianlai Sun, Ziyan Liu, Maria Litvinova, Alessandro Vespignani, Marco Ajelli, Cécile Viboud, and Hongjie Yu. Transmission heterogeneities, kinetics, and controllability of sars-cov-2. *Science*, 371(6526), 2021.

17. Ramanan Laxminarayan, Brian Wahl, Shankar Reddy Dudala, K. Gopal, Chandra Mohan B, S. Neelima, K. S. Jawahar Reddy, J. Radhakrishnan, and Joseph A. Lewnard. Epidemiology and transmission dynamics of covid-19 in two indian states. *Science*, 370(6517):691–697, 2020.

18. Tom Britton, Frank Ball, and Pieter Trapman. A mathematical model reveals the influence of population heterogeneity on herd immunity to sars-cov-2. *Science*, 2020.

19. Robert M. May and Roy M. Anderson. Spatial heterogeneity and the design of immunization programs. *Mathematical Biosciences*, 72(1):83 – 111, 1984.

20. Roy M Anderson and Robert M May. *Infectious diseases of humans: dynamics and control*. Oxford university press, 1992.

21. Stephen C. Hadler, Maria Alcala de Monzon, Dalia Rivero Lugo, and Maira Perez. Effect of timing of hepatitis b vaccine doses on response to vaccine in yucpa indians. *Vaccine*, 7(2):106 – 110, 1989. Special Conference Issue.
